# Digital health technologies and machine learning augment patient reported outcomes to remotely characterise rheumatoid arthritis

**DOI:** 10.1101/2022.11.18.22282305

**Authors:** Andrew P. Creagh, Valentin Hamy, Hang Yuan, Gert Mertes, Ryan Tomlinson, Wen-Hung Chen, Rachel Williams, Christopher Llop, Christopher Yee, Mei Sheng Duh, Aiden Doherty, Luis Garcia-Gancedo, David A. Clifton

## Abstract

Digital measures of health status captured during daily life could greatly augment current in-clinic assessments for rheumatoid arthritis (RA), to enable better assessment of disease progression and impact. This work presents results from weaRAble-PRO, a 14-day observational study, which aimed to investigate how digital health technologies (DHT), such as smartphones and wearables, could augment patient reported outcomes (PRO) to determine RA status and severity in a study of 30 moderate-to-severe RA patients, compared to 30 matched healthy controls (HC). Sensor-based measures of health status, mobility, dexterity, fatigue, and other RA specific symptoms were extracted from daily iPhone guided tests (GT), as well as actigraphy and heart rate sensor data, which was passively recorded from patients’ Apple smartwatch continuously over the study duration. We subsequently developed a machine learning (ML) framework to distinguish RA status and to estimate RA severity. It was found that daily wearable sensor-outcomes robustly distinguished RA from HC participants (F1, 0.807). Furthermore, by day 7 of the study (half-way), a sufficient volume of data had been collected to reliably capture the characteristics of RA participants. In addition, we observed that the detection of RA severity levels could be improved by augmenting standard patient reported outcomes with sensor-based features (F1, 0.833) in comparison to using PRO assessments alone (F1, 0.759), and that the combination of modalities could reliability measure continuous RA severity, as determined by the clinician-assessed RAPID-3 score at baseline (*r*^2^, 0.692; RMSE, 1.33). The ability to measure the impact of disease during daily life—through objective and remote digital outcomes—paves the way forward to enable the development of more patient-centric and personalised measurements for use in RA clinical trials.

## 1 Introduction

Rheumatoid arthritis (RA) patients follow subtle and unpredictable disease courses, patient-to-patient, with a progressive decline in physical function and quality of life and over time— often leading to disability and difficulty to perform many tasks of daily life^1^. RA symptoms include joint pain or tenderness, joint swelling, morning stiffness, reduction in joint range of movement (ROM), muscle pain, and fatigue^1^. Currently, the gold-standard methods to measure the impact of RA on daily life rely on infrequent clinical visits that may often occur every 3–4 months, with assessments depending on a combination of subjective clinician-determined scores^2^ and patient-reported outcomes^3^. These have inherent limitations, however, in that they can be subjective and are prone to recall bias^4,5^. As such, there is a need to objectively measure the impact of RA on daily life^6^, remotely over a continuous period, rather than restricting assessments to only intermittent physician visits. In recent years, consumer-grade mobile applications (app.) and wearable devices have shown promise to objectively measure participants’ symptoms during daily life^7^; these digital health technologies (DHT) tools^8^ have shown to increase study engagement, improve patient convenience, streamline collection of PROs^9^, and potentially generate more frequent and accurate data that can characterise disease^10^. DHT have been shown to measure RA symptoms and functions, such as range of motion (ROM) and gait-specific metrics during prescribed “active” assessments^11,12^. Other studies have shown how “passive” wearable actigraphy sensor-outcome measurements capture differences in RA physical activity (PA) in daily life, compared to healthy controls (HC)^13^, as well as to detect flaring of RA symptoms^14^.

However, there remains a lack of sufficient evidence for how DHT can provide objective insights into the impact of therapies for RA, despite progress made in other disease areas^15–22^. Particularly, the benefit of sensor-outcomes generated from prescribed active assessments compared with passive monitoring has not yet been explored together. While digitised patient-reported outcomes (PROs) enable a patient the ability to regularly record their “subjective” experience of disease activity in remote settings^23^, it remains unclear how “objective” sensor-outcomes could provide additional insights that can augment PROs to better characterise the impact of RA on daily life. As part of this characterisation, the sensitivity of DHT to measure RA symptoms, such as the volume of remote data required and the number of sensor-outcome measurements needed, will also need to be determined. Finally, the application of DHT sensor-outcomes to monitor RA during daily life remains yet to be validated against standard in-clinic administered assessments of RA impact^24^.

In this study, we therefore aimed to investigate how active and passive sensor-based measurements should be combined using machine learning (ML) to distinguish RA status from healthy controls, to augment traditional patient self-reported outcome (PRO) data, and to estimate standard in-clinic assessments of RA severity. Our work offers the first comprehensive evaluation of how sensor data captured during daily life can characterise RA status and severity, which represents an important first step towards the development of more sensitive and patient-centric measurements for use in RA clinical trials and real-world studies.

In order to investigate the objectives of this study, we performed the following set of analysis and experiments. We first illustrate the variety of sensor-based measurements that can be extracted from daily prescribed (active) smartphone-based assessments and (passive) smartwatch-based activity monitoring in an RA cohort. In this, we evaluate how smartwatch-based daily physical activity patterns can be remotely estimated using our bespoke deep convolutional neural, pre-trained using multi-task self-supervised learning (SSL) on a large-scale open-source cohort. We next assess the ability of our sensor-based measurements to identify RA status from healthy controls and to distinguish RA severity levels. As part of our analysis, we also explore the volume of days and number of sensor-outcomes required to remotely distinguish RA status. Finally, we investigated the power of active and passive sensor-outcomes to augment routinely collected patient self-reported outcome (PRO) data to estimate RA severity—as measured by standard in-clinic assessments of RA, such as the RAPID-3^25^.

## 2 Results

The GSK weaRAble-PRO study (GSK212295) was a 14-day observational study which investigated how DHT tools could objectively measure the impact of RA on participants’ daily lives. Digital wearable devices—a wrist-worn Apple Watch for passive monitoring and an iPhone, integrated with a bespoke mobile app. which prescribed daily guided assessments—collected high-frequency, objective sensor data in 30 RA patients and 30 matched Healthy Controls (HCs). Figure 1 provides an illustrative overview of the objectives of this study. Sensor-based measures of physical function, mobility, dexterity, and other RA specific symptoms were extracted from daily prescribed (active) iPhone guided tests using a combination of bespoke algorithms and proprietary algorithms developed by Apple ResearchKit, for instance, a wrist-range of motion exercise, a walking assessment, a nine-hole peg test, as well as two pose transition-based mobility exercises, lie-to-stand (LTS) and sit-to-stand (STS). In addition, continuous (passive) actigraphy was recorded from participants’ Apple smartwatch over the study duration in order to characterise daily activity patterns and sleep. In order to illustrate the various characteristics of RA we are interested in assessing, we have grouped measurements in Fig. 1 into four domains: physical function, daytime activity, daily living, and sleep; denoting particular types of measurements which may attribute to each domain. Note: this manuscript details a sub-study of weaRAble-PRO; trial design, feasibility, participant adherence, and other primary related study outcomes will be published as part of a complementary manuscript. Two RA participants withdrew immediately after enrolling in the study. Data from these participants were not collected, leaving 28 RA participants, 28 matched HCs, and 2 unmatched HCs for a total of 58 participant

**Figure 1.**
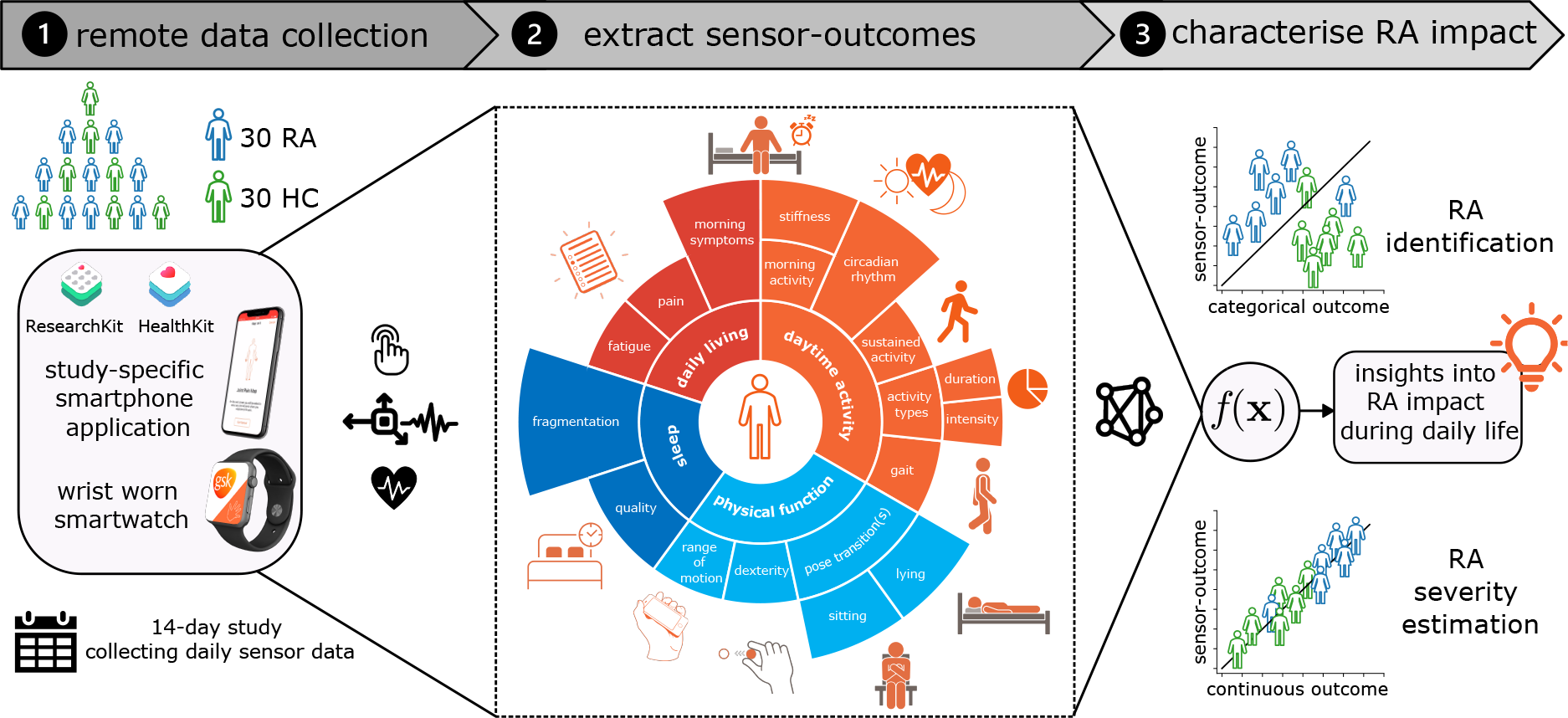
Illustration detailing the objectives of this study. The weaRAble-PRO 14-day trial aimed to investigate how digital health technologies (DHT)—a wrist-worn Apple smartwatch and an iPhone device, with bespoke mobile apps.—could augment patient reported outcomes (PRO) to characterise the impact of rheumatoid arthritis (RA) during the daily life of 30 moderate-to-severe RA patients, compared to 30 matched healthy controls (HC). We explore the ability of machine learning (ML) models to (1) estimate categorical RA outcomes, such as identifying RA participants from healthy controls and (2) estimate continuous RA outcomes, such as RA severity, using a combination of PRO and sensor-outcomes. Icon key: 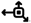, triaxial accelerometer + gyroscope sensor; 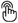, touch screen sensor; 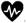, heart rate sensor; 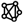, machine learning model.

### 2.1 Assessing smartwatch-based daily physical activity patterns

The daily physical activity of RA participants and healthy controls were estimated with a deep convolutional neural network (DCNN) that was first pre-trained on 100,000 participants in the publicly available UK Biobank, following a multi-task self-supervised learning (SSL) methodology^26^, which was subsequently fine-tuned on the free-living Capture-24 dataset^27^ of *<*150 participants to determine broad activity patterns of interest {sleep, sedentary, light physical activity, moderate-to-vigorous physical activity (MVPA)}^28,29^ and fine-grained activity prediction labels {sleep, sitting/standing, mixed, vehicle, walking, bicycling}^27^. In this study, we build upon our previous work by adding a temporal dependency to the DCNN (SSL) through a hidden markov model (HMM), which was appended to obtain a more accurate sequence of predicted activities over the continuous study period. It was found that the DCNN (SSL) + HMM improved broad activity estimation in Capture-24 (*κ*, 0.862 ± 0.088; F1, 0.815 ± 0.103) as compared to a baseline random forest (RF) + HMM approach (*κ*, 0.813 ± 0.108; F1, 0.775 ± 0.117)^27^. Next, the fine-tuned DCNN (SSL) + HMM model transformed the raw Apple smartwatch sensor data in weaRAble-PRO to determine participants’ daily activity patterns over the 14-day study period, for example, the time spent walking, the frequency of exercise, the length and quality of sleep, and other RA-specific measures, such as morning stiffness. Activity predictions were qualitatively evaluated over the entire RA and HC study population and demonstrated excellent face validity (see section A and section B for additional details).

### 2.2 Analysis of sensor-outcomes to distinguish RA status and severity levels

The raw smartphone and smartwatch data recorded during the (active) guided test exercises, and passively during the participants’ daily life, respectively, were summarised as sensor-outcome features. Univariate analysis demonstrated that a total of 153 (93%) sensor-based features (passive, n=131 (94%); active, n=22 (88%)) displayed significantly different medians (after post-hoc correction for multiple comparisons) between HC and RA severity groups (Kruskal–Wallis H test, p*<*0.05). A further 47 (34%) passive features, compared to 6 (24%) active features, were also significantly different (Mann-Whitney U test, p*<*0.05) between healthy and RA participants. Figure 2 compares the (fortnightly) average feature distributions between healthy controls (HC), RA (moderate) and RA (severe) participants for a selection of examples of passively collected smartwatch features (Fig. 2a–2c) and active guided test sensor features (Fig. 2d–2f) and a selection of patient self-reported outcomes recorded on the smartphone application (Fig. 2g–2i).

**Figure 2.**
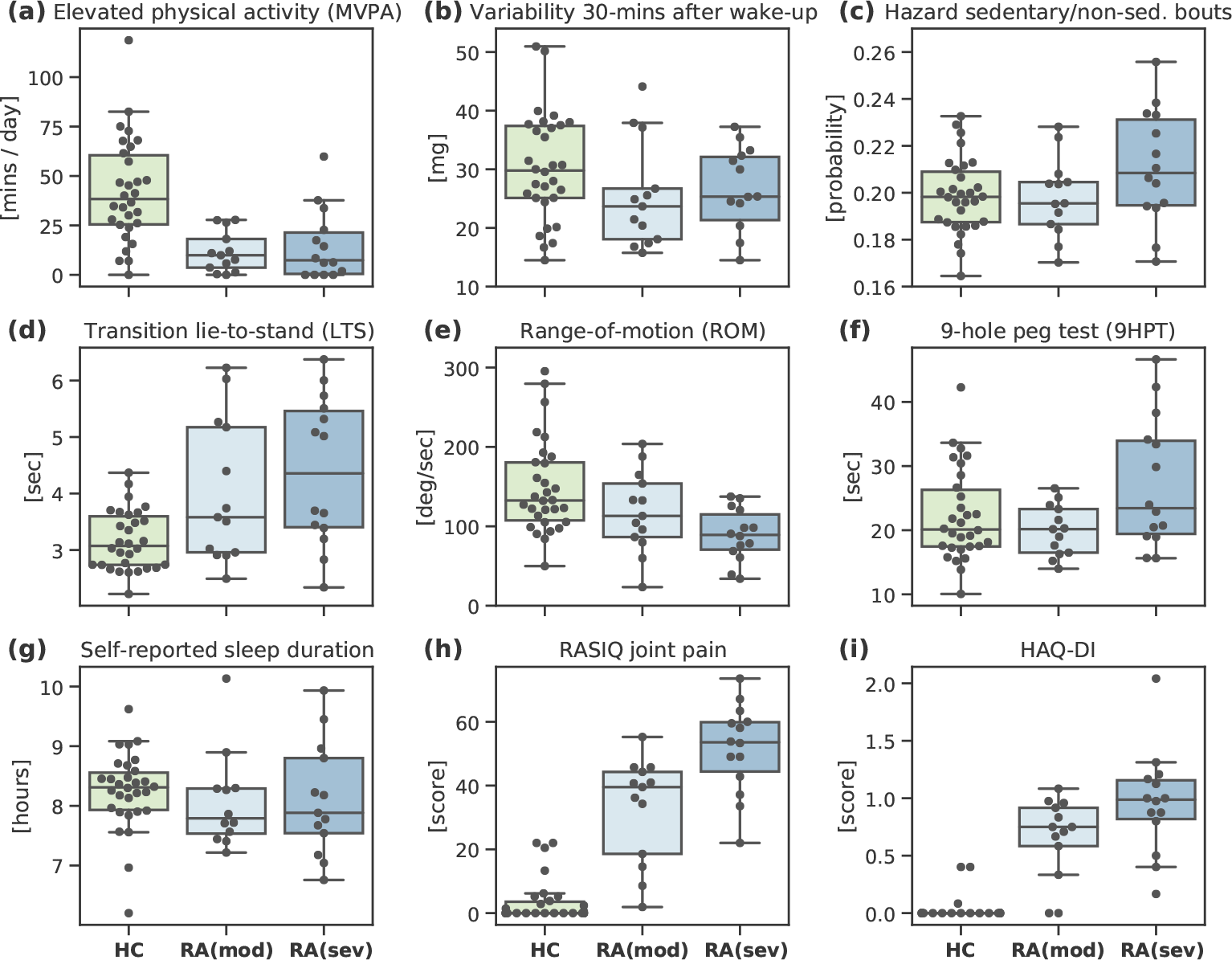
Ability of individual sensor-outcomes to distinguish between RA status and RA severity levels. Comparison of the average feature distributions per participants, between healthy controls (HC), RA (moderate) and RA (severe) groups for: (a–c) selection of passively collected smartwatch features; (d–f) selection of guided test collected smartphone features; and (g–i) selection of patient self-reported outcomes recorded on the smartphone application. For all examples shown, medians were significantly different between HC and RA groups: One-way ANOVA determined from the Kruskal–Wallis H-test, p*<*0.001. Abbreviations: deg, degrees; HAQ-DI, Health Assessment Questionnaire-Disability Index; mins, minutes; mg, mili-gravity acceleration units; MVPA, moderate-to-vigorous physical activity; RASIQ, GSK RA symptom and impact questionnaire; sed, sedentary; sec, seconds.

In order to explore the ability of many wearable sensor-outcomes to distinguish symptoms of RA from otherwise healthy individuals, and therefore measure the impact of RA during daily life, we devised a number of multivariate classification-based experiments. First, we investigated the performance of regularised logistic regression (LR) to differentiate RA participants from healthy controls using both passively collected activity monitoring features and guided test exercise features. Comparing model performance between sources (Fig. 3a), passive activity monitoring-based sensor features better distinguished RA participants using fortnightly averaged features (F1, 0.786) versus active (guided test) features (F1, 0.778). It was found that 12 subjects were misclassified using active-only models and 12 for passive-only, with just 4/12 (33%) of the same subjects incorrectly identified by both sources, 3 of which were the same HC participants. Combining active and passive wearable sensor features yielded in the highest performing models to distinguish RA participants overall, for example, using fortnightly averaged features from both sources (F1, 0.807) (for further expansion of results, see supplementary table A.I). It should be also be noted that linear logistic regression was found to perform comparatively to non-linear ensembles of decision trees, a Random Forest (RF) model and Extreme Gradient Boosted Trees (XGB)—as such this work subsequently opted to explore simple linear models for further analysis (see supplementary material, table A.II for expansion of results).

**Figure 3.**
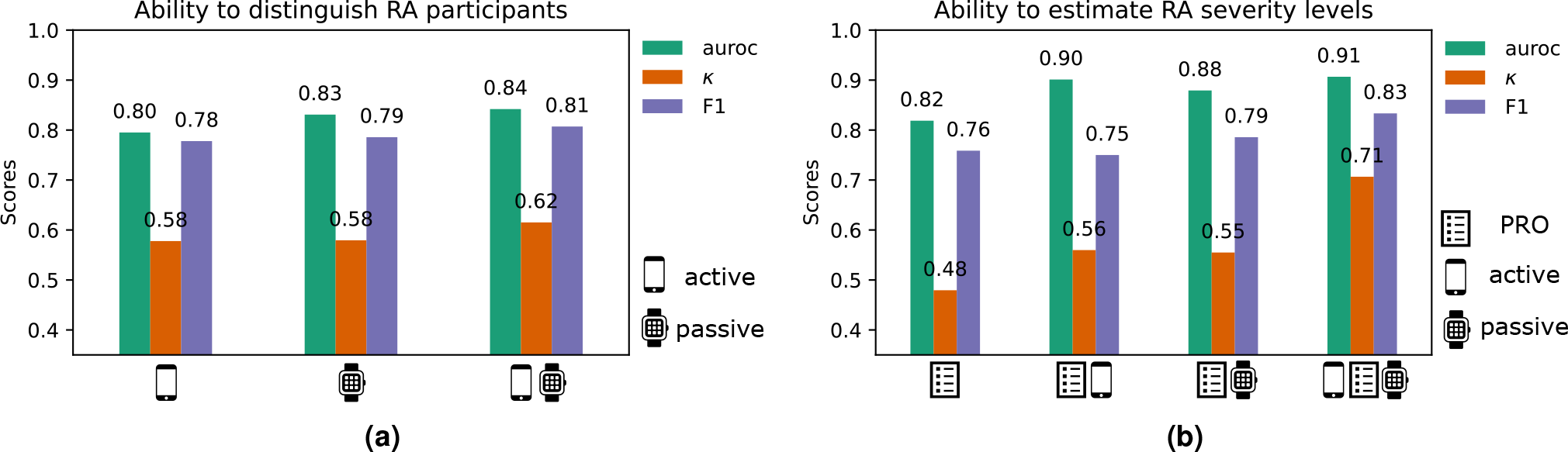
Ability of combined sensor-outcomes to distinguish between RA status and RA severity levels. Comparison of (a) RA identification (RA vs. HC) performance and (b) RA severity level estimation (RA (mod) vs RA (sev)), using patient reported outcomes (PRO) and combined PRO, active, and passive sensor-based outcomes in the weaRAble-PRO study. auroc: Area under the receiver operator curve; *κ*, Cohen’s Kappa statistic; F_1_, Macro-F1 score.

This study next investigated the ability of multiple sensor-based outcomes to augment PRO data in order to stratify RA severity levels. In weaRAble-PRO, participants were denoted as having moderate or severe RA based on baseline clinician-assessed RAPID-3 scores. Following similar procedure to RA identification, LR regularised models were investigated in order to distinguish RA (mod) and RA (sev) as binary classification tasks using fortnightly averaged study data. The benefit of incorporating additional sensor-based outcomes to patient (self-) reported outcomes is presented in Fig. 3b (expanded in supplementary table A.IV). It was observed that the linear combination of PRO assessments could accurately stratify RA symptom severity (F1, 0.759). The fusion of PRO data and sensor-based outcomes improved RA severity level estimation further with the addition of active (F1, 0.750) or passive (F1, 0.786) sources. Finally, the amalgamation of PRO outcomes with both active and passive sensor-based outcomes resulted in the most accurate RA severity level estimation (F1, 0.833)—an improvement of 10% compared to PRO outcomes alone (Fig. 3b). For further information on the selected PRO + sensor-outcomes, we refer the reader to supplementary table A.V.

### 2.3 Estimating the volume of days and number of sensor-outcomes required to remotely distinguish RA status

In weaRAble-PRO, participants performed daily guided test exercises—resulting in daily sensor features—and continuously recorded Apple Watch sensor data were summarised as daily activity monitoring-based features, over the 14-day study period. In this work, we aimed to determine the minimal number of days of sensor data required build a stable and robust estimate of disease status in RA participants compared to HC over the 14-day study period. Figure 4a represents an experiment exploring the (observation-wise) out-of-sample RA classification performance as a function of varying the number of non-contiguous days of data that are averaged per participant. Evaluated over 500 randomly sampled permutations of non-contiguous days, results (median + IQR) indicated that RA prediction stabilised once more than 7 non-contiguous days of data were used per participant. Furthermore, we found that averaging daily feature values over weekly and fortnightly periods improved model performance. However, it was observed that model performance using weekly averaged features was often similar to fortnightly averaged (we refer the reader to supplementary table A.I for further details).

**Figure 4.**
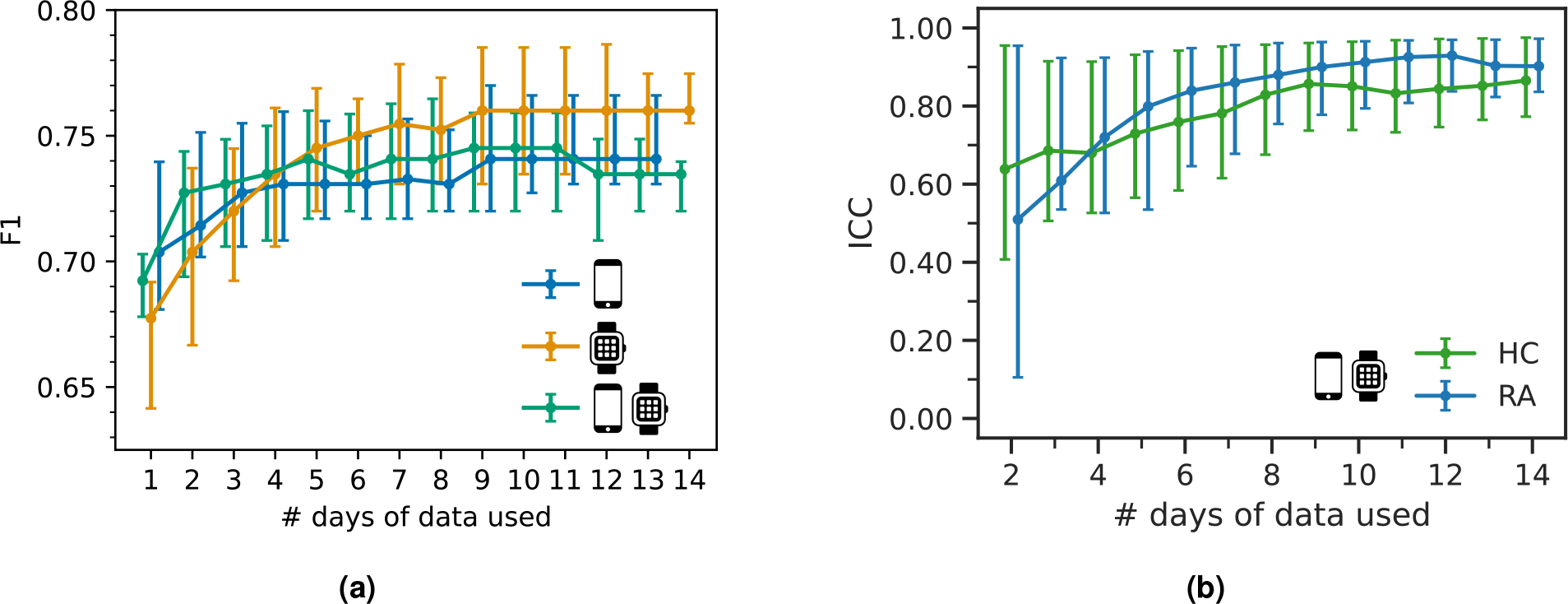
The number of days of sensor-data required to remotely characterise RA impact. Comparison of **(a)** the minimal amount of days of data needed distinguish RA status, as measured by the F1 score across 5-fold cross validation (CV), between active, passive, and combined feature sources; **(b)** the feature (test-retest) reliability, as measured by the intraclass correlation coefficient (ICC), between RA participants and HC across the study duration (14 days); F1 scores and ICCs suggest that model performance and feature reliability stabilises once more than 7 days of data are used per participant.

To investigate feature consistency and reproducibility, the intra-class correlation coefficient (ICC) for each feature was evaluated over the study duration (14 days). ICCs were calculated for each feature using *n* = [2, 3, …, 14] days of data per participant, individually for HC and RA participants. Higher ICC’s suggest a high degree of similarity on the performance of each task over the course of the study, and lower coefficients mean that participants tended to perform the task differently each day of the study. ICC’s for HCs ranged from 0.582 to 0.854, while those for RA participants ranged from 0.424 to 0.897. Figure 4b depicts the median + inter-quartile range (IQR) of ICC values for the LR-elastic net retained active + passive features. Intra-rater reliability analyses suggest that feature reliability stabilises to good (ICC=0.75–0.9) and excellent (ICC*>*0.9) once more than 7 contiguous days of data were used per participant.

In order to evaluate the number of sensor-outcomes required to remotely distinguish RA status, we compared various feature regularisation techniques, lasso (*𝓁*_1_), ridge (*𝓁*_2_), elastic-net (*𝓁*_1_+*𝓁*_2_), and sparse-group lasso, using fortnightly (i.e., study duration) averaged features. It was found that introducing sparsity through regularisation improved classification performance. In addition, active and passively recorded sensor-based features could be grouped into domains, based on the guided test they were extracted from, or the perceived functional domain of daily activity they were assumed to assess. Introducing group-wise sparsity with the sparse-group lasso (SG-lasso), regularising on the number of groups (i.e., the feature domains) and the coefficients within each group, resulted in the highest RA participant identification performance (F1, 0.807), compared to lasso (*𝓁*_1_, F1, 0.772), ridge (*𝓁*_2_, F1, 0.792), and elastic net (*𝓁*_1_+*𝓁*_2_, F1, 0.792) regularisation (for expansion of results, see supplementary material, table A.II). The features and groups selected by each regularisation technique are illustrated in Fig. 5, represented as the mean LR coefficient value **w** over CV per each feature and feature domain (coefficient values have been normalised between 0–1 to benefit comparison between models). Examining the feature sparsity of elastic-net (*𝓁*_1_ + *𝓁*_2_) (Fig. 5a), it was observed that features from multiple domains were selected. In contrast, the SG-lasso, as shown in Fig. 5b, selected mostly passive activity-based smartwatch features—TVDA with some morning stiffness measures—to distinguish RA status. Group sparsity penalised simultaneously selecting from multiple feature domains, where within group-sparsity regularised the feature coefficient values within the selected domains. Using fewer domains and features, the SG-lasso was able achieve similar performance to LR elastic-net, even marginally improving performance (F1, 0.807). For further details on the features extracted, and selected, we refer the reader to the supplementary material sections A and E respectively.

**Figure 5.**
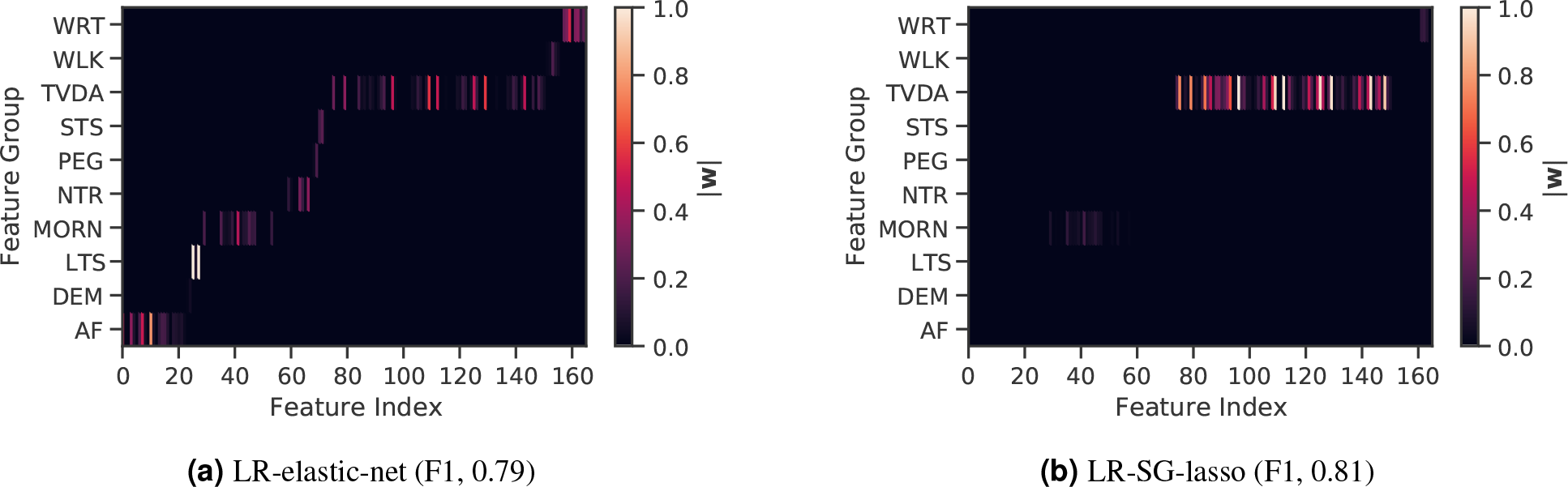
The number of sensor-outcomes required to remotely distinguish RA status. Comparison of features selected between regularised logistic regression (LR) models for: (a) elastic-net (F1, 0.79) and (b) SG-lasso (F1, 0.81). The SG-lasso promotes group-wise sparsity (i.e., regularising the number of feature domains) and within-group sparsity (i.e., regularising the number of features per domain), achieving a similar performance to LR elastic-net, while selecting a fewer number of domains and features. Feature importance, denoted as the mean LR coefficient value (**w**) over cross validation, are illustrated by colour intensity. Sensor-based feature domain abbreviations: AF: activity fragmentation; DEM: demographics; LTS: lie-to-stand assessment; MORN: morning stiffness; NTR: night-time restlessness; PEG, 9-hole peg test; STS: sit-to-stand assessment; TVDA: total volume of daytime activity; WLK: walking assessment; WRT: wrist assessment.

### 2.4 Estimating in-clinic RA severity scores from PRO and sensor-based outcomes

Rheumatoid arthritis severity levels were denoted by a clinician administered RAPID-3 assessment^25^ at baseline in the weaRAble-PRO study. The RAPID-3—a “rapid” and easy to administer questionnaire—is also validated against more exhaustive assessments for RA, such as the disease activity score 28 (DAS28) and clinical disease activity index (CDAI) in clinical trials and clinical care^25^. In this work, we aimed to establish how the combination of PRO and sensor-based outcomes could stratify continuous RAPID-3 RA severity. Note: HC subjects were assigned a RAPID-3 score of zero at baseline. Through multivariate modelling, using LR elastic-net, it was determined that PRO and sensor-based features could accurately estimate RAPID-3 scores to within 1 point (*r*^2^, 0.69; MAE, 0.94; RMSE, 1.33), an improvement compared to using PRO measures alone (*r*^2^, 0.63; MAE, 1.16; RMSE, 1.45). The association between actual and PRO + sensor-outcome estimated RAPID-3 scores was found to be good-to-excellent (*r >*0.75), Pearson’s r=0.60, p*<*0.001; Spearman’s *ρ*=0.83, p*<*0.001.

Participants in weaRAble-PRO were also administered a twice-daily interactive Joint Pain Map (JMAP) questionnaire on their iPhone^11^, in order to more precisely record and localise perceived pain. Participant model-estimated RAPID-3 scores were further interpreted through detailed inspection of the daily smartphone-based patient-reported joint pain map (JMAP) total scores—an external validation measure, which was not included as a predictor in the model—as expanded in Fig. 6. The JMAP score, defined as the sum of all individual joint pain scores per recording, was intended as a coarse measure to holistically capture participants’ overall level of perceived pain, in addition to validated PRO assessments. Higher JMAP scores indicate higher levels of pain experienced. It was observed that RAPID-3 estimations were reliable and robust, in that they faithfully characterised RA participant’s perceived level of symptoms, through the JMAP. For example, in Fig. 6, the RA (sev.) participant with consistently the largest reported degree of pain across the 14-day study exhibited the highest actual RAPID-3 score (6.7), which was closely estimated by the model as 7.1. JMAP scores further enabled additional explanation of model performance, especially with respect to RAPID-3 estimations that were not reflective of actual RAPID-3 scores. For instance, the RA (mod) participant with the lowest estimated RAPID-3 score (0.2) actually reported zero pain experienced over the 14-day study duration, despite a RAPID-3 assignment of 3.7 at baseline. Non-zero estimated RAPID-3 scores for some HC could also often be contextualised, due to these participants frequently self-reporting low-levels of pain in their JMAP (i.e., non-zero JMAP entries) over the study period, despite being healthy. As such, it was determined that PRO and sensor-based RAPID-3 estimates reliably reflected participant’s RA symptoms over the study.

**Figure 6.**
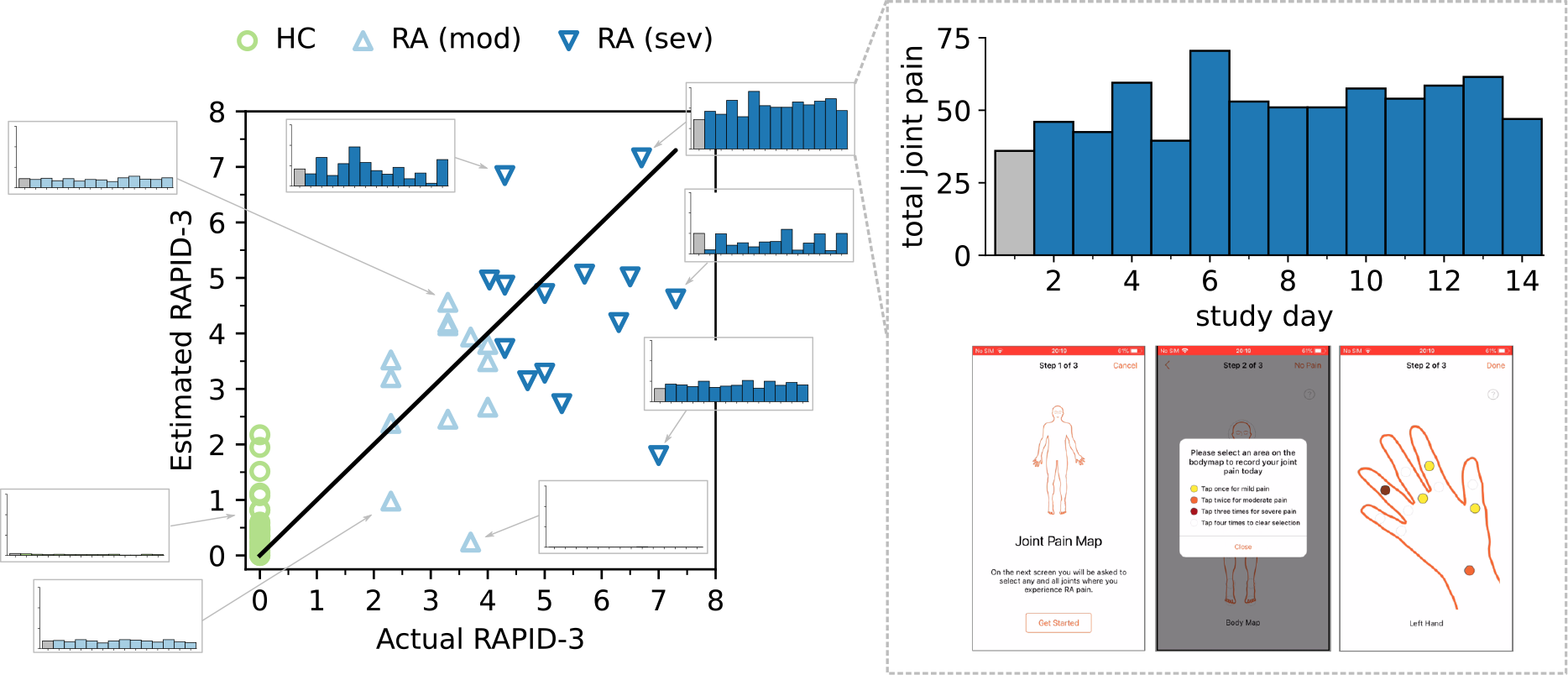
The ability of remote PRO + sensor-outcomes to estimate in-clinic determined RA severity scores. Scatter plot of baseline RAPID-3 scores *y* versus predicted *ŷ* scores per subject, using elastic net with PRO + sensor-outcomes, over cross-validation (CV). Participant model-estimated RAPID-3 scores can be further interpreted through detailed inspection of the daily smartphone-based patient-reported joint pain map (JMAP) total scores—which was not included as a predictor in the model. Higher JMAP scores indicate higher levels of pain experienced. Additional interpretability, through the JMAP, demonstrated that PRO + sensor-based outcome estimation of the RAPID-3 could reliably reflect patient’s perceived daily RA symptoms. Note: Baseline JMAP total scores, recorded on the same day as the baseline RAPID-3, are denoted in grey; the JMAP y-axis scale is the same among all subplots. HC subjects were assigned a RAPID-3 score of zero at baseline. A black line represents perfect predictions (*r*^2^, 0.692; MAE, 0.938; RMSE, 1.333).

## 3 Discussion

Our findings in the weaRAble-PRO study demonstrate how digital health technology (DHT) captured sensor-outcomes, recorded from smartphone-based active tests, and continuously collected passive smartwatch-based monitoring, could characterise meaningful aspects of rheumatoid arthritis (RA) impairment and physical function impacting daily life. Remotely collected wearable sensor-outcomes could distinguish RA status from healthy controls—demonstrating further improved performance when combining the sensor-data from both devices—and how objective sensor-outcomes could augment patient (self-) reported outcomes to remotely estimate RA severity. Furthermore, by the half-way point of the weaRAble-PRO study (day 7), a sufficient volume of data had already been collected to reliably distinguish the characteristics of RA participants. This work provides the first comprehensive evaluation how remote and objective digital sensor-outcomes enrich our ability to understand the impact of RA on daily life between clinical visits.

In this work, we detailed how raw data collected from smartphone and smartwatch sensors can be transformed into sensor-based outcomes that are reflective of disease status. In concurrence with previous studies, many remotely collected smartphone sensor-outcomes distinguished RA participants and RA severity levels. For example, it was observed that joint ROM features differentiated HC and RA groups—a similar finding to our previous work^12^—and that RA participants were less mobile, taking longer to move between positions (as measured during the lie-to-stand exercise)—as previously shown by Andreu-Perez, et al.^30^. Continuously collected smartwatch sensor data, known as passive monitoring, allowed the measurement of aspects of RA daily life, such as physical activity, sleep, and other RA specific symptoms, such as morning stiffness, or night-time restlessness. In this study we trained an activity recognition model on the free-living Capture-24 dataset to estimate daily activity patterns in the weaRAble-PRO population. Leveraging the latest advances in self-supervised learning (SSL) allowed our model to be pretrained on 100,000 participants with 700,000 days of diverse, unlabelled wearable sensor data in the UK Biobank^26^, which combined with HMM temporal smoothing, significantly improved activity prediction compared to our previous established RF-HMM based methods^27,29^. Our SSL DCNN+HMM model enabled a more robust and fine-grained estimation of daily activity patterns beyond traditional acceleration magnitude levels^13,14^, which we proposed could allow a richer characterisation of PA and sleep in RA. Activity monitoring revealed distinct differences distinguishing RA status, for example the daily percent of the day in moderate-to-vigorous physical activity, and similar features, were significantly lower in the RA population compared to healthy controls—a similar finding by Prioreschi, et al.^13^, and an observation people with RA regularly self-report^31^. Other specific RA symptom measurements, like morning stiffness or disrupted sleep, were evident in certain RA participants. For example, the mean acceleration value *>* 30 [mins] after wake-up were lower in RA—also a similar finding to Keogh, et al.^32^—or that the number of movement episodes during night-time sleep distinguished some specific RA participants. We also observed that after collecting 7 days of sensor-data in the weaRAble-PRO study, a sufficient volume of data had already been recorded to reliably distinguish RA participants from a healthy population; participant feature reliability (as measured ICC values) stabilised at good-to-excellent levels, maximal identification performance of RA participants plateaued, and that there was no additional benefit to averaging over a fortnight’s worth of data versus a week. Therefore it is recommended that considering at least one week’s worth of sensor data is collected, it might be more beneficial to gather less data from a greater number of participants, rather than greater duration of sensor data from the same participants.

Our work is the first study to combine active smartphone and passive wearable measurements to distinguish RA status and measure variations in RA severity. While models trained on only passive features tended to marginally outperform models trained solely on active guided test features, combining both active + passive features led to the best performance in RA identification for all models investigated. Interestingly, it was found that different subjects were misclassified by active versus passive models. For example, 12 subjects were misclassified using active-only models and 12 for passive-only, with just 4/12 (33%) of the same subjects incorrectly identified by both sources, 3 of which were the same HC participants. In addition, further experiments with the LR-SG-lasso determined that only activity monitoring domain features were mainly needed in order to distinguish RA participants from health controls. This indicates that we sometimes do not need to prescribe all guided test assessments, or to parse all activity feature domains, but that a small number of prescribed assessments can be sufficient to characterise RA status. For example, including only the lie-to-stand assessment rather than also prescribing the similar, and highly correlated, sit-to-stand assessment in future studies; or removing the prescribed walking assessment (shown to have little predictive value in the weaRAble-PRO study), and using passive daily life walking predictions generated from the activity recognition model instead, which could reduce patient burden. Finally, we also found that combining patient-reported outcomes (PRO) and objective sensor-outcomes could better capture RAPID-3-based RA severity at baseline than PROs alone; most estimated RAPID-3 scores correctly stratified participants across severity levels from healthy to moderate to severe RA, suggesting that sufficient information to characterise RA disease severity could be reflected in the remote monitoring outcomes derived in the 14-day weaRAble-PRO study. To the best of the authors knowledge, this offers the first evaluation and insight how remote monitoring outcomes in daily life can estimate in-clinic administered assessments of RA impact.

There are a number of limitations that must be considered in the weaRAble-PRO study. Despite rich individual level measurements, the study recruited a relatively small sample size (HC, n=30; RA, n=30). As such, a degree of variability and uncertainty existed in constructing cross-validated models to distinguish RA participants, RA severity levels, or estimate the in-clinic RAPID-3 assessment. Extrapolation of results aimed at generalising RA is therefore not possible without the availability of larger cohorts and further external validation. In addition, this study only recruited RA patients with moderate-to-severe levels of disease activity; future studies should also aim to characterise patients with lower levels of disease activity or those in remission. There were also limitations associated with modelling a clinician-administered assessment, or clinical labels formulated from in-clinic assessments. For instance, the RAPID-3 was assessed at baseline, with participants recalling the prior week, yet the PRO and sensor-based features were calculated as averages over subsequent 14-day trial period from baseline. As such, the baseline RAPID-3 may not have precisely reflected the participant’s disease status recorded earlier, due to the underlying mutability and heterogeneity of RA symptoms over short periods of time. The subjectivity of PRO predictors should also considered, for instance, pain or perceived quality of sleep is relative, and some healthy participants recorded experiencing pain or affected sleep in PRO questionnaires. As a result, some PRO values influenced HC RAPID-3 predictions greater than zero, i.e., indicating the presence of RA symptoms—albeit non-zero estimated RAPID-3 predictions for HCs were generally low (*<*2).

The weaRAble-PRO study typifies how continuously collected patient self-reported and sensor-based outcomes may more closely reflect participant perceived and experienced symptoms that impact daily life. While in-clinic assessments are considered the gold-standard means of assessing disease severity in RA, it is clear that remotely collected, continuous, patient-centric measurements generated from PRO and sensor-based outcomes offer promising insights that can undoubtedly augment in-clinic assessments for RA. We believe that our work—the first comprehensive evaluation how remote sensor data can augment traditional PRO measures to estimate clinician-determined RA severity—helps informs future DHT study design to better characterise the impact of RA on daily life, ultimately to expand the use of DHT to develop more sensitive, and patient-centric, endpoints in RA clinical trials and real-world studies.

## 4 Methods

### 4.1 Dataset

Remotely collected smartphone and smartwatch sensor data was obtained from the GSK study title: Novel Digital Technologies for the Assessment of Objective Measures and Patient Reported Outcomes in Rheumatoid Arthritis Patients: A Pilot Study Using a Wrist-Worn Device and Bespoke Mobile App. (212295, weaRAble-PRO). This observational study followed 30 participants diagnosed with moderate-to-severe RA and 30 matched HCs over 14 days. The population demographics, in-clinic, and relevant patient self-reported outcomes, as assessed at baseline, are reported in table 1. RA participants were denoted as displaying moderate disability, RA (mod), or severe disability, RA (sev), as determined by their baseline RAPID-3 score. Note: Two RA participants withdrew immediately after enrolling in the study. Data from these participants were not collected, leaving 28 RA participants, 28 matched HCs, and 2 unmatched HCs for a total of 58 participants. Further study details, including participant requirement and data collection, are outlined in the accompanying supplementary material.

**Table 1.** Population demographics, in-clinic, and selected patient self-reported outcomes, as assessed at baseline, where the mean ± standard deviation across the population are reported.

|  | HC<sup>a</sup><br>(n=28) | RA (mod)<sup>b</sup><br>(n=13) | RA (sev)<sup>c</sup><br>(n=15) | *p*<sup>1</sup> |
| --- | --- | --- | --- | --- |
| **Demographics** |  |  |  |  |
| Age, years | 58.4 ± 9.9 | 56.9 ± 11.4 | 60.4 ± 7.1 | 0.33 |
| Female, n (%) | 25 (89%) | 11 (84%) | 14 (93%) | 0.92 |
| BMI | 25.8 ± 4.6 | 31.1 ± 5.9 | 31.7 ± 8.6 | 0.96 |
| **In-clinic Outcome(s)** |  |  |  |  |
| RAPID-3 | 0 ± 0 | 3.2 ± 0.7 | 5.3 ± 1.1 | <0.001 |
| **Patient Reported Outcome(s)** |  |  |  |  |
| HAQ-DI | 0 ± 0 | 0.63 ± 0.36 | 1.03 ± 0.42 | <0.01 |
| RASIQ-pain | 3.1 ± 6.7 | 32.1 ± 20.8 | 56.2 ± 11.6 | <0.01 |
| RASIQ-stiffness | 5.9 ± 9.5 | 33.9 ± 18.9 | 51.6 ± 10.2 | <0.05 |
| RASIQ-impact | 47.3 ± 5.0 | 53.9 ± 5.1 | 50.8 ± 7.6 | 0.33 |
| FACIT | 49.2 ± 2.9 | 38.9 ± 4.3 | 31.9 ± 7.6 | <0.05 |
| PROMIS-sleep | 49.6 ± 2.8 | 52.7 ± 4.2 | 52.4 ± 4.3 | 0.83 |
| PROMIS-pain | 42.2 ± 4.8 | 54.2 ± 7.29 | 58.8 ± 4.6 | 0.09 |
| JMAP total pain<sup>2</sup> | 0.20 ± 0.5 | 13.5 ± 13.9 | 18.8 ± 13.7 | 0.23 |

<sup>1</sup> *p*, *p*-value calculated from Mann Whitney U-test comparing severe vs. moderate RA participants;
<sup>2</sup> Note: self-reported JMAP is not a validated PRO in RA;
<sup>a</sup> Matched HC to RA participants only;
<sup>b</sup> RA participants with baseline RAPID-3: 6.1–12.
<sup>c</sup> RA participants with baseline RAPID-3: >12;

#### Sensor-based data collection

The Apple Watch and iPhone were used to collect high frequency raw sensor data from predefined, (active) guided tests on a daily basis. Participants were prescribed daily to perform five iPhone-based assessments: WRT, a wrist range of motion (ROM) exercise^12^; WLK, a 30 second walking exercise^12^; PEG, a digital 9-hole peg test^33^; STS, a sit-to-stand transition exercise^30,34^; and LTS, a lie-to-stand transition exercise^30,34^. For more details on the (active) guided test sensor-based features extracted, see supplementary material C. A brief overview of the guided tests prescribed in weaRAble-PRO are presented in supplementary material section C.1. In addition, the Apple Watch was used to continuously collect background sensor data (denoted passive data), as the participants went about their daily activities. Participants were asked to maintain a charge on both the Apple Watch and the iPhone, so that interruptions to monitoring and data transfer were kept to a minimum. Since night-time activity was also monitored, while participants were asleep, it was requested that charging should be done during the day, in a way that fit the participants’ schedules (e.g., charging in the morning while getting ready for the day). For more details on the activity monitoring features, see supplementary material section B.5.

#### Patient-reported outcomes

Patient-reported outcomes (PRO), most often self-report questionnaires, were administered to assess disease activity, symptoms, and health status and quality of life from the patients’ perspective^35,36^. The weaRAble-PRO study administered a selection of validated PRO measures for RA in compliment to bespoke digital PRO assessments—that are validated in clinical trials, where the questions, response options, and the general approach to assessment were standardised for all participants. PROs were recorded on days 1, 7, and 14 of data collection. The PRO assessments administered to participants are outlined in supplementary material section D.3.

### 4.2 Smartwatch-based estimation of daily life patterns

In order to generate unobtrusive measures characterising physical activity and sleep in RA participants during daily life, the raw Apple Watch actigraphy (i.e., accelerometer) sensor data was transformed through a human activity recognition (HAR) sensor processing and deep convolutional neural network (DCNN) pipeline. Figure 7 illustrates how a deep convolutional neural network (DCNN) can transform raw Apple smartwatch sensor data to estimate a participant’s daily activity patterns in the weaRAble-PRO study using self-supervised learning (SSL). The construction of this pipeline yielded unobtrusively measured summary features of physical activity and sleep for RA participants, computed daily during normal life.

**Figure 7.**
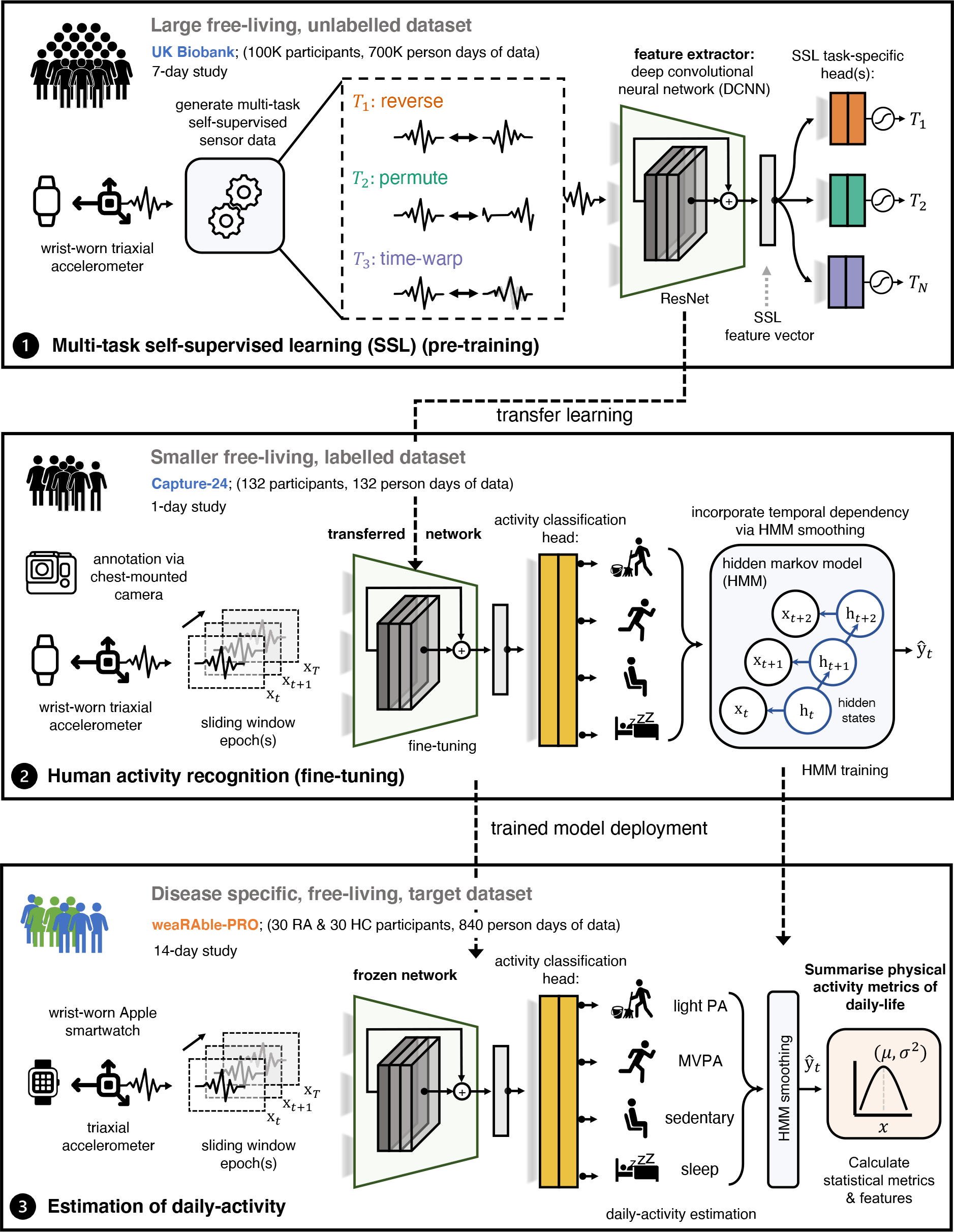
Self-supervised learning pipeline. Continuous (passive) actigraphy was recorded from patients’ Apple smartwatch over the study duration. Deep convolutional neural networks (DCNN) were pre-trained on 700,000 person days in the publicly available UK Biobank using self-supervised learning—and fine-tuned with the Capture-24 dataset—to estimate participant’s daily activity patterns in the weaRAble-PRO study. Physical activity (PA) metrics of daily-life, for example, the time spent walking, the frequency of exercise, or the length and quality of sleep were investigated as markers to characterise symptoms of disease in people with RA compared to HC.

A deep convolutional neural network (DCNN) with a ResNet-V2 architecture was first pre-trained following a multitask self-supervised learning (SSL) methodology on 100,000 participants—each participant contributing 7 days yielding roughly 700,000 person days of data—in the open-source UK biobank^26^. The SSL pre-trained model was then fine-tuned to perform activity recognition as a downstream task in the Capture-24 dataset.

The Capture-24 study is a manually labelled, free-living dataset—that is reflective of real-world environments—and is available for training an activity recognition model to be applied to the weaRAble-PRO study. In Capture-24, actigraphy data was collected for 24-hours from 132 healthy volunteer participants with a Axivity AX3 wrist-worn device as they went their normal day. Activity labels provided by photographs automatically captured roughly every 30 seconds by a wearable camera for each participant. Capture-24 was labelled with 213 activity labels, standardised from the compendium of physical activities^28^. Activity labels were then summarised into a small number of free-living behaviour labels, defining activity classes in Capture-24. There are two major labelling conventions used within Capture-24 that the model was trained to predict, defined as broad activity: {sleep, sedentary, light physical activity, moderate-to-vigorous physical activity (MVPA)}^28,29^; and fine-grained activity: {sleep, sitting/standing, mixed, vehicle, walking, bicycling}^27^.

HAR model predictions are essentially independent— meaning that the sequence of activities over each 30 second epoch incorporates no temporal information epoch-to-epoch, for instance how the previous epoch prediction affects the current, or next, activity prediction. In order to add temporal dependency to the HAR SSL model, a Hidden Markov Model (HMM) was implemented in a post-processing step to obtain a more accurate sequence of predicted activities over the continuous 14-day data collection period as per Willetts, et al.^27^.

The Capture-24 fine-tuned HAR SSL-HMM model was then implemented to estimate daily activities in weaRAble-PRO study data. For additional information of the HAR deep network, SSL, and other related information, we refer the reader to our previous work^26^. Further results relating to the SSL-HMM model are outlined in the supplementary material. The sensor processing pipeline developed for the Apple Watch in the weaRAble-PRO study is outlined in supplementary Fig. B.I and within the accompanying supplementary material.

### 4.3 Extraction of sensor-based outcomes

Wearable sensor-based features were derived from the smartphone during the active guided tasks and passively from the smartwatch during daily life. “Active” features, extracted from smartphone sensor-based measurements during the prescribed guided tests, aimed to capture specific aspects of RA physical function, related to pain, dexterity, mobility and fatigue^12^. In addition “passive” features were extracted from smartwatch sensor-based measurements, collected continuously in the background over the 14-day period. Daily activity predictions from the ML SSL model were summarised into general features measuring activity levels, period, duration and type of activity, as well as sleep detection and sleeping patterns. Furthermore, devised under the guidance of Rheumatologists, additional activity monitoring features specifically aimed at characterising well-known RA symptoms were also developed, such as morning stiffness and night-time restlessness.

The supplementary material sections B and C also detail algorithms used to extract active and passive features in the weaRAble-PRO study. For a full list of extracted sensor-based features in weaRAble-PRO, we refer the reader to the supplementary material table E.I.

### 4.4 Statistical Analysis

#### Univariate Testing

Pair-wise differences groups between groups, for example HC vs. RA, or RA (mod) vs. RA (sev) were analysed for the equality in population median using the non-parametric Mann-Whitney U test (MWUT)^37–39^. One-way analysis of variance (ANOVA) tests were also used to assess differences between medians of multiple groups, for example HC vs. RA (mod) vs. RA (sev) were assessed using the Kruskal-Wallis (KWt) test by ranks^40^. The Brown-Forsythe (BF) test by (absolute deviation) of medians was used to investigate if various groups of data have been drawn with equal variances^41^.

#### Correlation Analysis

Correlation analysis was utilised to determine the association or dependence between sets of random variables, such as the dependence between features, or for assessing a features’ clinical utility by measuring the association to an established clinical metric. This study investigated the (linear) Pearson’s *r* correlation and the (non-linear) Spearman’s Rho *ρ* correlation between features, between features and PROs, and between clinical assessments to determine levels of association. The strengths of the correlations were classified as good-to-excellent (r*>*0.75), moderate-to-good (*r*=0.50–0.75), fair (*r*=0.25–0.49) or no correlation (*r <*0.25)^42^.

#### Feature Reliability

Intra-rater (i.e., test-retest) reliability was determined using intra-class correlation coefficient (ICC) values^43^, which were used to assesses the degree of similarity between repeated features over the course of the study for each patient. In this work, the *ICC*(3, *k*) was calculated^44^–which considers the two-way random average measures with *k* repeated measurements—for the 14-day session across subjects, where the raters *k* are the study days. Reliability was categorised as either poor (ICC*<*0.5), moderate (ICC=0.5–0.75), good (ICC=0.75–0.9) or excellent (ICC*>*0.9)^45^.

#### Correcting for multiple hypothesis testing

Multiple hypothesis testing was performed due to the large volume of features by controlling the false discovery rate (FDR) at level *α* using the linear step-up procedure introduced by Benjamini and Hochberg (BH)^46,47^.

### 4.5 Machine-learning estimation of RA status and severity

This work explored how state-of-the art machine learning (ML) models characterise the impact of RA during the daily life of participants in over the 14-day weaRAble-PRO study. Multivariate modelling aimed to explore the ability of active, passive, and PRO measures to (1) distinguish RA participants from healthy controls (HC), and (2) to estimate RA disease severity: between RA participants with moderate symptoms (RA mod) and severe symptoms (RA sev) as binary classification tasks. Expansions of this analysis subsequently investigate how the in-clinic RAPID-3 assessment, a continuous measure of RA severity, could be estimated from the combination of PRO and sensor-based outcomes.

#### Overview of models

This analysis compared both linear and non-linear ML models to transform PRO and sensor-based outcomes to capture RA status and severity. Regularised linear regression (LR) models, with combinations of *𝓁*_1_ and *𝓁*_2_ priors, such as LR-lasso (*𝓁*_1_), LR-ridge (*𝓁*_2_), and LR-elasticnet (*𝓁*_1_ +*𝓁*_2_) were compared to yield predictive, yet sparse model solutions^48^. Further regularisation extensions were also investigated using the sparse-group lasso (SG-lasso)–an extension of the lasso that promotes both group sparsity and within group parameter-wise (*𝓁*_2_) sparsity, through a group lasso penalty and the lasso penalty—which aims to yield a sparse set of groups and also a sparse set of covariates in each selected group^49,50^

Linear regression regularised models were also compared to decision tree (DT) based non-linear models, for instance the off-the-shelf Random Forest (RF)^51^ and Extreme Gradient Boosted Trees (XGB)^52^. Both LR- and DT-based models can intrinsically perform regression or classification depending on the task required. In the LR case, classification is denoted as logistic regression (though a logit-link function). NOTE: in this analysis LR can refer to both linear regression for continuous outputs or logistic regression for classification outputs. In the DT case, the mean prediction of the individual trees creates a continuous output for regression. For further details on the models employed in this study, we refer the reader to the supplementary material section F.2.

### Model evaluation

To determine the generalisability of our models, a stratified leave-k-subjects out cross-validation (CV) was employed. This consisted of randomly partitioning the dataset into folds with k=5 subjects in each fold, which was stratified with equal class proportions where possible. Subject data remained independent between training, validation, and testing sets. One set was denoted the training set (in-sample), and the remaining 20% of the dataset was then denoted testing set (out-of-sample) on which predictions were made.

### Feature-wise and prediction-wise aggregation

In this work, we experimented with feature-wise and prediction-wise aggregation. In feature-wise aggregation, features were computed either as: daily feature values over the 14-day study period; the average daily feature value over a 7-day period (weekly); the average daily feature value over a 14-day period (fortnightly). Predictions could then be evaluated for each day (denoted *observation-wise*) or aggregated over all days through majority voting each individual prediction per subject (denoted *subject-wise*). For example, daily and weekly averaged features result in daily, or weekly predictions (i.e., *observation-wise*), which were summarised into *subject-wise* outcomes by majority voting over the repeated predictions.

### Evaluation metrics

Multi-class classification metrics were reported as the *observation-wise* median and interquartile (IQR) range over one CV, as well as the *subject-wise* outcome for that CV, using: auroc, area under the receiver operating characteristic curve; *k*, Cohen’s kappa statistic^53,54^; F_1_, F1-score. The coefficient of determination, *r*^2^, the mean absolute error (MAE), and root-mean squared error (RMSE) were used to evaluate modelling the (continuous) in-clinic RAPID-3 scores.^55^.

## Data Availability

Anonymised individual participant data that support the findings of this study are available from the corresponding author, upon reasonable request and subject to GSK's approval.

## Acknowledgements

The authors are grateful to all the study participants and their families for their time and dedication to this study. The authors would also like to thank Priyanka Bobbili PhD, Julien Bendelac BSc, Jessica Landry MSc, Maral DerSarkissian PhD, Mihran Yenikomshian MBA, and Med Kouaici (MEng) from Analysis Group (MA, USA) for their support in app. design & development and data collection, and to Elinor Mody from Reliant Medical Group (MA, USA) for patient recruitment. The weaRAble-PRO study was funded and sponsored by GSK Plc. The research described in this paper was funded by GSK Plc. Authors conducting this research also acknowledge support by the National Institute for Health Research (NIHR) Oxford Biomedical Research Centre (BRC). Aiden Doherty is supported by the Wellcome Trust [223100/Z/21/Z].

## Code Availability

Apple Watch sensor processing was performed using a bespoke version of the biobankAccelerometerAnalysis toolkit, found at: https://github.com/OxWearables/biobankAccelerometerAnalysis. Deep networks were built using Python v3.7 through a PyTorch v1.7 framework. Our self-supervised learning activity prediction code and trained models are publicly available at: https://github.com/OxWearables/ssl-wearables, including pre-trained models on 100K participants in the UK Biobank. Some guided test exercises and health metrics calculated are proprietary to Apple ResearchKit (http://researchkit.org/) and Apple HealthKit (https://developer.apple.com/documentation/healthkit) which we refer the reader for more details. Statistical and machine learning analysis was developed using scikit-learn v1.1.1. Further analysis code can be made available from the corresponding author upon reasonable request.

## Ethics Statement

All study information, informed consent, study questions and instructions for conducting the guided tests were first drafted in the form of a survey instrument. The survey instrument was then programmed into the mobile app. All documentation including the study protocol, any amendments, and informed consent procedures, were reviewed and approved by Reliant Medical Group’s IRB. All participants provided written informed consent before any study procedures were undertaken. The study was conducted in accordance with the International Committee for Harmonisation principles of Good Clinical Practice and the Declaration of Helsinki.

## Author Contributions

A.P.C conceptualised the data analysis, designed methodology, software and interpretation. V.H, H.Y, and G.M contributed software application for analysis. V.H, R.T, W-H.C, R.W, L.G-G contributed to the design of the study and towards the data analysis and interpretation. C.L, C.Y, M.S.D were involved in the design of the study, data collection, and software for data acquisition. A.D, L.G-G, D.A.C jointly supervised. A.P.C wrote the manuscript; all other authors: review & editing.

## A Supplementary Results

### A.1 Extended Results: Assessing smartwatch-based daily physical activity patterns

Figure A.Ia–A.Ib summarises the population-wide daily activity by time of the day for HC and RA groups. The probability of an activity being performed at a specific time can be computed as the number of instances detected for that activity across all data points (participants and days) at that time divided by the total number of data points. Representative examples of the predicted daily activity patterns for an individual healthy control (HC) and RA (moderate) participant are depicted in Fig. A.Ic–A.Id respectively. The times when the Apple Watch was left to charge can be clearly seen in each example, indicated by the white non-wear times, typically occurring after wake-up or before bedtime. Both participants demonstrated consistent wake-up and bed times, day-to-day—which the activity prediction model tended to correctly identify.

### A.2 Extended Results: Multivariate feature analysis

The relationship between the wearable sensor-based features extracted in this study, for both active (smartphone) and passive (smartwatch) data sources were investigated using pair-wise Spearman’s *ρ* correlation. Correlation analysis indicated good-to-excellent relationships (*ρ >* 0.75) between many features within feature domains (intra-source); for example, most TVDA features were highly correlated with each other (positively and negatively). Analysis also revealed good-to-excellent (*ρ >* 0.75) correlation between domains of features sources (inter-source); for example, TVDA features were not only correlated with each other, but with other passive feature domains, such as AF or MORN features. Much of the inter-source correlation was between similar domains, such as within the activity monitoring-based feature domains, or within the guided test (active) feature domains—suggesting a high degree of multicollinearity and redundancy. However, mostly fair correlations (*ρ*=0.25–0.50) between active and passively extracted sensor features suggested that different information may be learned during activity monitoring versus guided test exercises. The resulting correlation matrix is depicted in Fig. A.II.

**Figure A.I.**
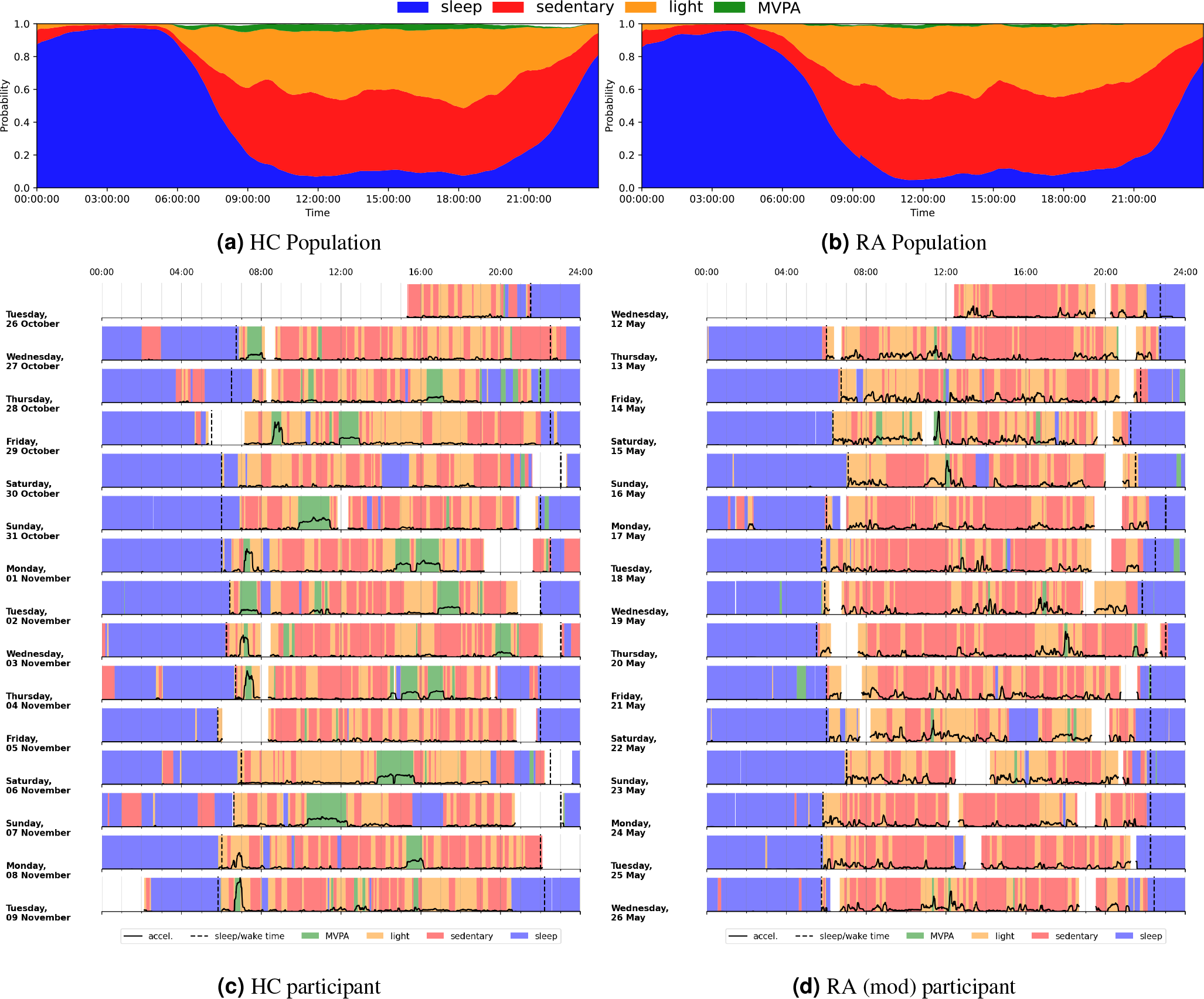
Assessing smartwatch-based daily physical activity patterns. Variation in the average predicted daily-activity (probability) over time for all (a) HC participants and (b) RA participants in the 14-day weaRAble-PRO study. Predicted daily activity patterns for an individual (c) healthy control (HC) participant, (Female, 66 yrs.) and (d) a moderate Rheumatoid Arthritis (RA mod) participant (Female, 50 yrs.; RAPID-3, 3.7). Moving average acceleration values are overlaid in black. Participant self-reported sleep / wake times are indicated with long-dashed black lines. Non-wear times (expected daily for watch charging) are indicated by white areas. Note: the acceleration y-axis scaling between (c) and (d) is not the same due to difference in the magnitude of acceleration between participants. MVPA, moderate-to-vigorous physical activity.

**Figure A.II.**
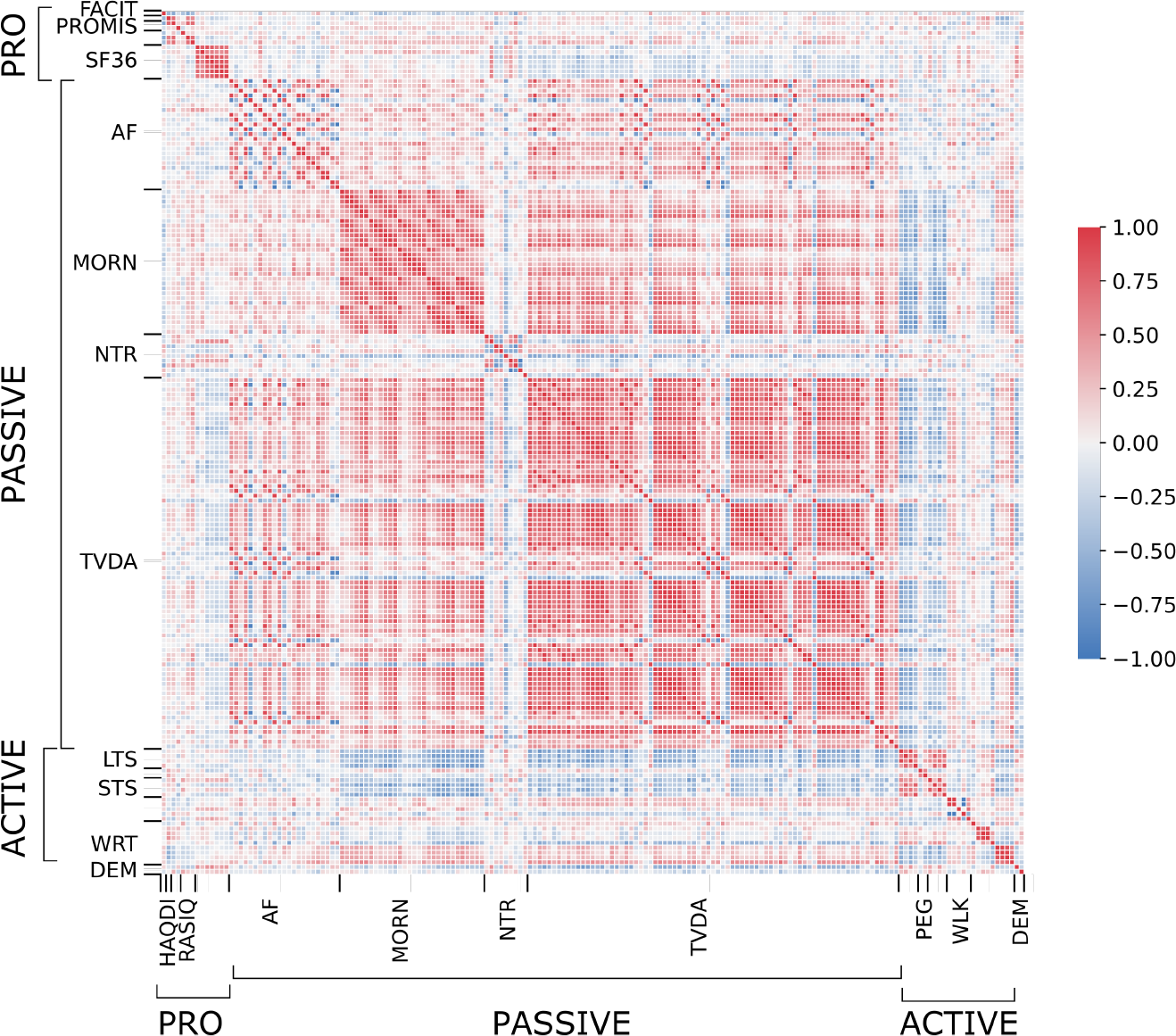
Assessing correlation and collinearity between PRO and sensor-based features. Pairwise Spearman’s *ρ* correlation matrix for PRO, active, and passive features, labelled by feature domain. Feature association is bounded between +1/-1 denoting positive and negative correlation. Feature domain abbreviations: FACIT: Functional Assessment of Chronic Illness Therapy Fatigue; HAQ-DI: Health Assessment Questionnaire-Disability Index; PROMIS: Patient-Reported Outcomes Measurement Information System; RASIQ: GSK RA symptom and impact questionnaire; SF-36: Short-Form 36 (SF-36); AF: activity fragmentation; MORN: morning stiffness; NTR: night-time restlessness; TVDA: total volume of daytime activity; LTS: lie-to-stand assessment; PEG: 9-hole peg test; STS: sit-to-stand assessment; WRT: wrist assessment; WLK: walking assessment; DEM: demographics.

### A.3 Extended Results: Distinguishing RA participants

Table A.III represents a selection of features that were retained by LR-elastic-net. The model tended to pick features from all domains, but consistently tended to select many different features between cross validation splits. Some features however, for example, the mean transition time [sec] from standing to lying, daily average total time in MVPA bouts [mins], the average hazard of non-MVPA to MVPA bouts were constantly chosen over all data splits. Other features were selected less often but, when chosen, weighted highly in the model, for instance: median ROM [deg] or the number of movement episodes during night-time sleep [count/hr].

**Table A.I.** Comparison of RA vs. HC classification performance across different source and feature combinations with 5-fold cross-validation (CV). Results are presented as: (1) the posterior overall *subject-wise* outcome for one cross-validation (CV) run as well as (2) the *observation-wise* median and inter-quartile range (IQR) across that CV in brackets. The best performing model for each source combination are highlighted in **bold**. auroc: Area under the receiver operator curve; *κ*, Cohen’s Kappa statistic; F_1_, Macro-F1 score.

| source | feature<sup>1</sup> | model | auroc | $\kappa$ | $F_1$ |
| --- | --- | --- | --- | --- | --- |
| active | daily | LR-elastic-net | 0.673 (0.681, 0.616–0.748) | 0.183 (0.180, 0.090–0.421) | 0.531 (0.648, 0.464–0.676) |
|  |  | LR-SG-lasso | 0.817 (0.725, 0.676–0.930) | 0.542 (0.433, 0.258–0.708) | 0.755 (0.680, 0.610–0.831) |
|  | weekly | LR-elastic-net | 0.699 (0.757, 0.629–0.792) | 0.362 (0.374, 0.118–0.459) | 0.640 (0.667, 0.444–0.700) |
|  |  | **LR-SG-lasso** | **0.802 (0.771, 0.729–0.825)** | **0.614 (0.471, 0.408–0.545)** | **0.800 (0.667, 0.667–0.762)** |
|  | fortnightly | LR-elastic-net | 0.779 (0.743, 0.686–0.800) | 0.333 (0.267, 0.098–0.633) | 0.655 (0.667, 0.545–0.800) |
|  |  | LR-SG-lasso | 0.795 (0.743, 0.714–0.833) | 0.578 (0.500, 0.471–0.633) | 0.778 (0.727, 0.667–0.800) |
| passive | daily | LR-elastic-net | 0.821 (0.691, 0.680–0.701) | 0.439 (0.347, 0.323–0.362) | 0.714 (0.689, 0.617–0.691) |
|  |  | LR-SG-lasso | 0.852 (0.712, 0.677–0.758) | 0.609 (0.369, 0.335–0.380) | 0.776 (0.691, 0.667–0.698) |
|  | weekly | LR-elastic-net | 0.837 (0.795, 0.722–0.859) | 0.579 (0.441, 0.438–0.507) | 0.786 (0.706, 0.667–0.733) |
|  |  | LR-SG-lasso | 0.812 (0.833, 0.804–0.906) | 0.612 (0.588, 0.571–0.607) | 0.795 (0.750, 0.714–0.828) |
|  | fortnightly | **LR-elastic-net** | **0.790 (0.800, 0.657–0.943)** | **0.615 (0.500, 0.500–0.814)** | **0.807 (0.727, 0.727–0.889)** |
|  |  | LR-SG-lasso | 0.831 (0.867, 0.657–0.943) | 0.579 (0.500, 0.314–0.814) | 0.786 (0.727, 0.714–0.889) |
| active + passive | daily | LR-elastic-net | 0.814 (0.729, 0.704–0.891) | 0.473 (0.312, 0.196–0.552) | 0.727 (0.686, 0.611–0.767) |
|  |  | LR-SG-lasso | 0.837 (0.744, 0.704–0.908) | 0.504 (0.317, 0.275–0.625) | 0.720 (0.683, 0.605–0.808) |
|  | weekly | LR-elastic-net | 0.848 (0.842, 0.786–0.850) | 0.614 (0.538, 0.486–0.577) | 0.800 (0.769, 0.762–0.800) |
|  |  | LR-SG-lasso | 0.848 (0.833, 0.700–0.893) | 0.614 (0.589, 0.254–0.814) | 0.800 (0.783, 0.692–0.889) |
|  | fortnightly | LR-elastic-net | 0.857 (0.914, 0.867–0.914) | 0.579 (0.471, 0.351–0.814) | 0.786 (0.714, 0.667–0.889) |
|  |  | **LR-SG-lasso** | **0.842 (0.867, 0.714–0.943)** | **0.615 (0.676, 0.500–0.814)** | **0.807 (0.833, 0.727–0.889)** |

<sup>1</sup> daily: daily feature values over the 14-day study period; weekly: the average daily feature value over a 7-day period; fortnightly: the average daily feature value over a 14-day period;

**Table A.II.** Comparison of RA vs. HC classification performance for logistic regression (LR) based models and decision trees (DT) across with 5-fold cross-validation (CV) with fortnightly (i.e., study duration) averaged active + passive features. Results are presented as: (1) the posterior overall *subject-wise* outcome for one cross-validation (CV) run as well as (2) the *observation-wise* median and inter-quartile range (IQR) across that CV in brackets. The best performing model for each feature representation are highlighted in **bold**. auroc: Area under the receiver operator curve; *κ*, Cohen’s Kappa statistic; F_1_, Macro-F1 score.

| | model | auroc | $\kappa$ | f1 |
| --- | --- | --- | --- | --- |
| LR | - | 0.853 (0.867, 0.800–0.943) | 0.542 (0.500, 0.459–0.814) | 0.755 (0.727, 0.727–0.889) |
|  | lasso | 0.788 (0.857, 0.667–0.867) | 0.545 (0.500, 0.241–0.814) | 0.772 (0.727, 0.714–0.889) |
|  | ridge | 0.777 (0.833, 0.767–0.914) | 0.612 (0.657, 0.441–0.814) | 0.792 (0.800, 0.769–0.889) |
|  | elastic-net | 0.801 (0.867, 0.667–0.914) | 0.612 (0.657, 0.441–0.814) | 0.792 (0.800, 0.769–0.889) |
|  | **SG-lasso** | **0.842 (0.867, 0.714–0.943)** | **0.615 (0.676, 0.500–0.814)** | **0.807 (0.833, 0.727–0.889)** |
| DT | RF | 0.862 (0.933, 0.829–0.957) | 0.646 (0.657, 0.471–0.814) | 0.800 (0.800, 0.727–0.889) |
|  | XGB | 0.851 (0.833, 0.829–0.914) | 0.614 (0.676, 0.471–0.814) | 0.800 (0.833, 0.714–0.889) |

In order to determine the test-retest reliability of the selected features, we calculated intra-class correlation coefficient (ICC) values^43^, which were used to assesses the degree of similarity between repeated features over the course of the study for each patient. Here, we calculated *ICC*(3, *k*)^44^–which considers the two-way random average measures with *k* repeated measurements—for the 14-day session across subjects, where the raters *k* are the study days.

The minimal detectable change, with a 95% confidence interval (CI) (MDC_95_), was also calculated to determine the minimal change in a feature which is greater than the within subject variability and measurement error, indicating how much a measured change is likely to reflect true change from repeated measurement. First, the standard error of measurement (SEM), which provides an absolute index of precision^42^ was calculated:

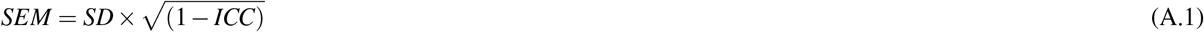

where *SD* and *ICC* are the variance and intra-class correlation coefficients of the feature, *x*. Next, the minimal detectable change, with a 95% confidence interval (CI) (MDC_95_) was determined^42,56^:

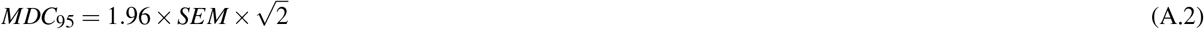

where *MDC*_95_ was expressed as percentages that are independent of the units of measurement for each feature:

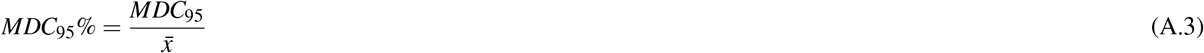

using the respective mean feature value, 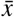.

**Table A.III.**
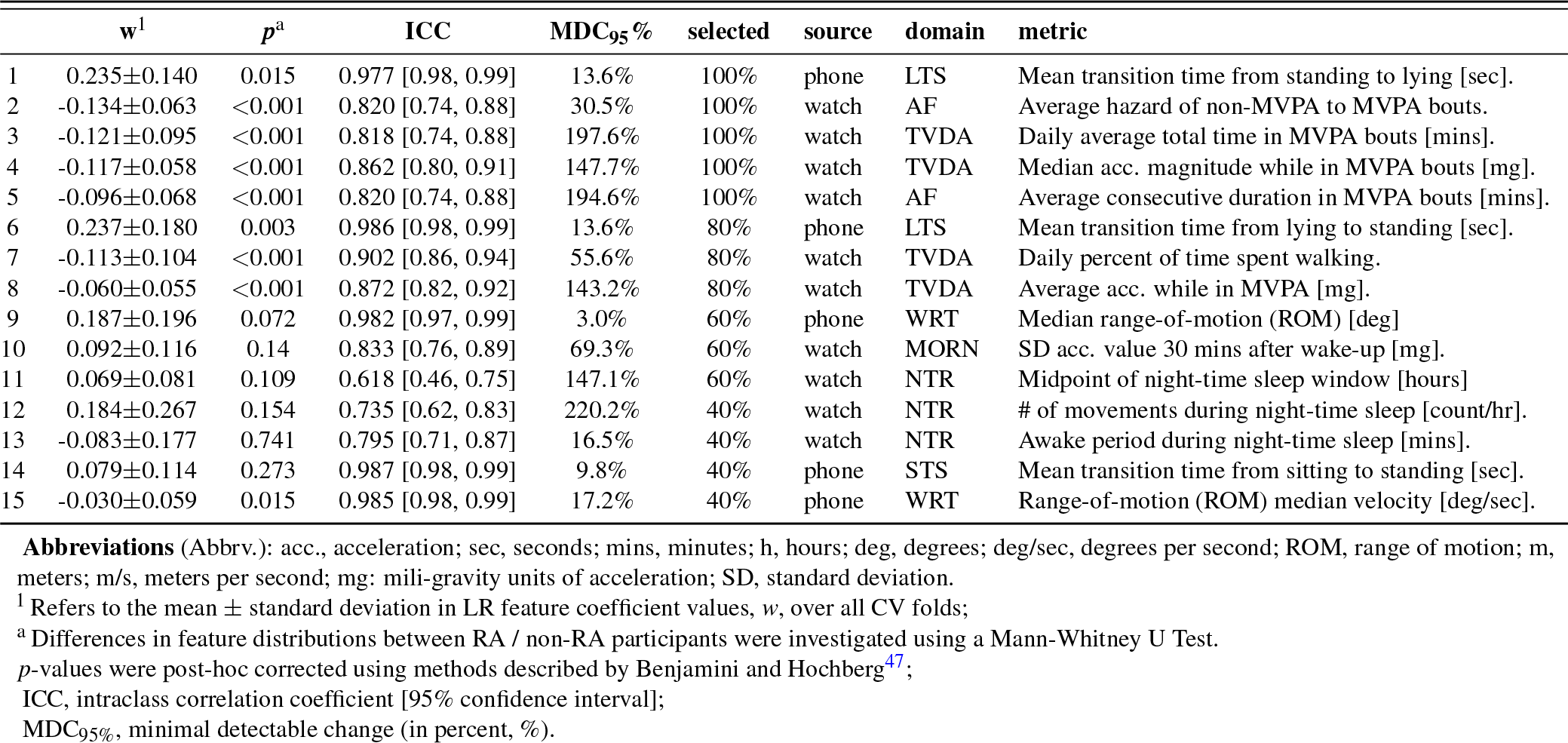
Selection of top performing active (smartphone) and passive (smartwatch) extracted features for RA identification, as determined by logistic regression (LR) elastic-net across 5-fold subject-wise cross validation (CV), with fortnightly (i.e., study duration) averaged features. Features were ranked per CV fold by increasing shrinkage regularisation parameter *λ* and recording the percentage (%) of time that feature is selected in the subset that minimises the CV error in the validation set. Feature domain abbreviations: AF: activity fragmentation; DEM: demographics; LTS: lie-to-stand assessment; MORN: morning stiffness; NTR: night-time restlessness; STS: sit-to-stand assessment; TVDA: total volume of daytime activity; WLK: walking assessment; WRT: wrist assessment.

### A.4 Extended Results: Distinguishing RA severity levels

**Table A.IV.**
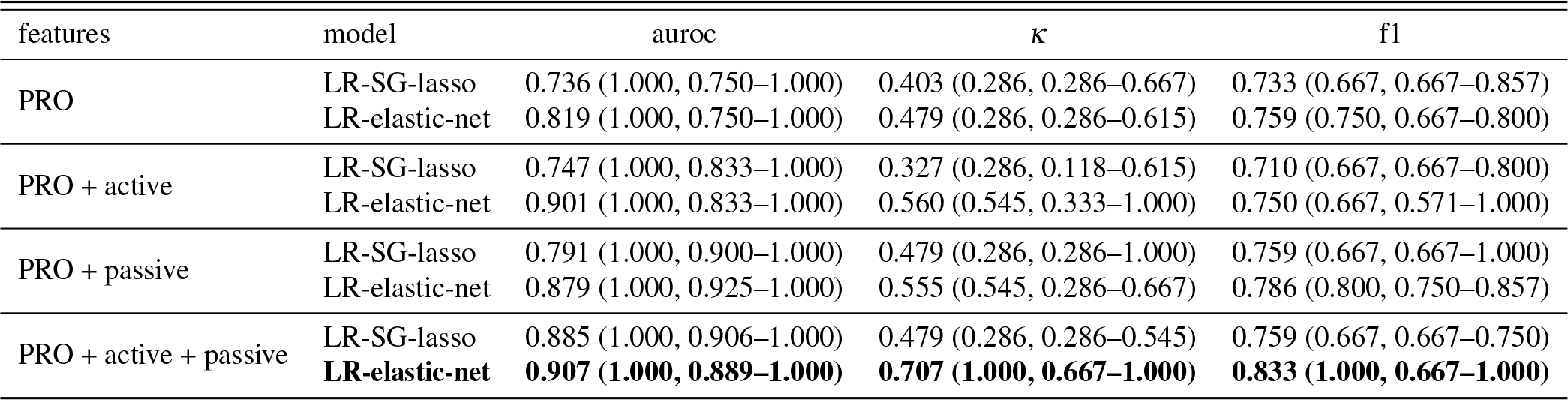
Comparison of RA severity level prediction using patient reported outcomes (PRO), versus using PRO + sensor-outcomes, over 5-fold cross-validation (CV) with fortnightly (i.e., study duration) averaged active + passive features. Results are presented as: (1) the posterior overall *subject-wise* outcome for one cross-validation (CV) run as well as (2) the *observation-wise* median and inter-quartile range (IQR) across that CV in brackets. The best performing model for each feature representation are highlighted in **bold**. auroc: Area under the receiver operator curve; *κ*, Cohen’s Kappa statistic; F_1_, Macro-F1 score.

**Table A.V.** Top 10 selected features from PRO + sensor-outcome based RA severity level estimation, as determined by LR-elastic-net across 5-fold subject-wise cross validation (CV), with fortnightly (i.e., study duration) averaged features.

| | $w^1$ | $p^a$ | selected | source | domain | metric |
| --- | --- | --- | --- | --- | --- | --- |
| 1 | 1.876±0.670 | 0.002 | 100% | PRO | RASIQ | Joint pain |
| 2 | 1.696±1.066 | 0.07 | 100% | phone | PEG | 9HPT total time [sec] |
| 3 | 1.324±0.762 | 0.45 | 100% | - | DEM | Age range [5 years] |
| 4 | 1.177±1.128 | 0.13 | 100% | phone | WRT | Range-of-motion (ROM) median velocity [deg/s] |
| 5 | 0.971±0.774 | 0.09 | 100% | watch | TVDA | # continuous periods of walking > 30 mins [count] |
| 6 | -0.410±0.276 | 0.53 | 100% | - | DEM | Sex [M/F] |
| 7 | -0.965±0.897 | 0.10 | 80% | phone | WRT | Range-of-motion (ROM) [deg] |
| 8 | 0.543±0.825 | 0.07 | 80% | PRO | RASIQ | Joint stiffness |
| 9 | 0.430±0.410 | 0.12 | 80% | watch | AF | Average hazard of sedentary to non-sedentary bouts |
| 10 | 0.904±0.887 | 0.02 | 60% | PRO | HAQ | HAQ-DI total score |

**Abbreviations** (Abbrev.): PRO, patient-reported outcome; DEM, demographics information; acc., acceleration; s, seconds; mins, minutes; h, hours; deg, degrees; deg/s, degrees per second; ROM, range of motion; m, meters; m/s, meters per second; mg: mili-gravity units of acceleration.
<sup>1</sup> Refers to the mean ± standard deviation in LR feature coefficient values, $w$ , over all CV folds;
<sup>a</sup> Differences in feature distributions between RA (mod) / RA (sev) participants were investigated using a Mann-Whitney U Test. $p$ -values were post-hoc corrected using methods described by Benjamini and Hochberg<sup>47</sup>;

## B Methodology: Smartwatch sensor feature extraction

### B.1 Sensor Processing Pipeline

The sensor processing pipeline developed for the Apple Watch in the weaRAble-PRO study is outlined in Fig. B.I, yielding unobtrusively measured summary features of physical activity and sleep for RA participants, computed daily during normal life.

**Figure B.I.**
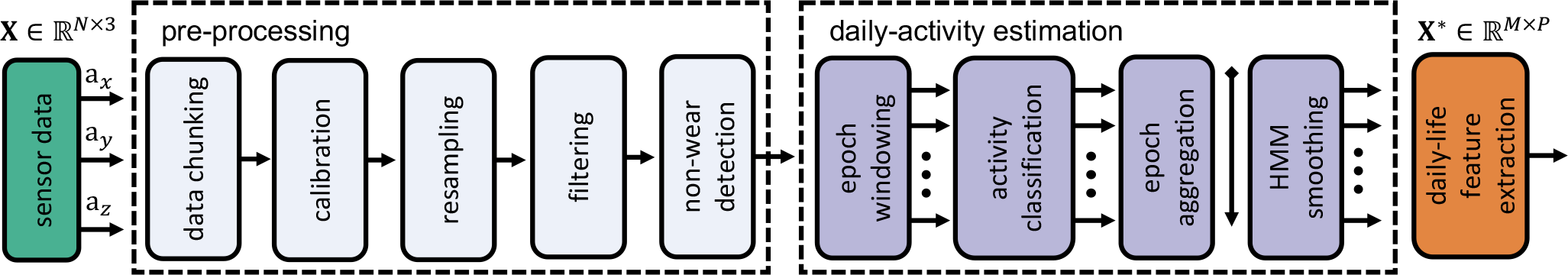
Sensor processing pipeline developed for the Apple Watch in the weaRAble-PRO study. The raw 3-axis accelerometer sensor data, (**a**_*x*_, **a**_*y*_, **a**_*z*_), denoted by **X** ∈ ℝ^*N×*3^, where *N* are the number of continuously collected accelerometer samples over the 14-day study period (*N* = 50 [*Hz*]×3600 [*sec*]×24 [*hr*]×14 [*days*]), can be transformed into measures characterising physical activity and sleep, **X**^*\**^ ∈ ℝ^*N×*3^, where *M* is the new sampling range, *daily* (*M* = 14), and *P* are the number of measures of daily life (i.e., features). In this case, a *N ≫M* problem has been reduced into useful *M ×P* features, unobtrusively measuring the physical activity and sleep of RA participants during daily life.

An overview of the pipeline is as follows:

1. **Pre-processing:**
  a. Data chunking, memory optimisation. Convert raw 50 Hz accelerometer data to daily chunks;
  b. Calibration to local gravity, local UTC timestamps;
  c. Resampling, 30 Hz;
  d. Butterworth, low-pass filtering at 17 Hz;
  e. Non-wear detection and segmentation;
2. **Daily-activity Estimation:**
  a. Epoch windowing, 30 [sec];
  b. Activity classification per epoch;
  c. Epoch aggregation, daily;
  d. Posterior activity prediction with hidden Markov model (HMM) smoothing, see section B.3;
3. **Characterising daily life:**
  a. Physical activity and sleep feature extraction, see section B.5 for more details;

### B.2 Deep Network-based Activity Recognition

In this work, a deep learning-based activity recognition model, known as a deep convolutional neural network (DCNN), was trained on Capture-24 and then used to directly estimate daily activity in the weaRAble-PRO study.

#### B.2.1 Multi-Task Self-Supervised Learning

Developing robust activity classification models is challenging in clinical studies due to the lack of labelled data for training. Deep networks, in particular, need a lot of training data in order to be robust and generaliseable. Open-source HAR-based datasets have small sample sizes, with generally n*<*100 participants as annotating free-living wearable data for human activity recognition (HAR) requires a concurrent video stream, and the labelling process is resource-intensive^26^.

There are however massive-scale unlabelled wearable datasets, such as the UK Biobank (UKB), which have collected data on roughly 100,000 participants with over 6 billion samples available. This study build upon our previous work demonstrating how advances in self-supervised learning (SSL) could help exploit the hidden information in these large-scale unlabelled datasets^26^. SSL consists of training a model on a pretext task in an unlabelled dataset (often in a multi-task problem). The supervised task is devised based on labels manually created in the unlabelled dataset, such as distinguishing transformed versus original data. The SSL model has to determine for each sample, if a transform has been applied or not, and what transform(s) have been applied as a multi-task problem. Essentially you create a robust deep feature extractor, built on a diverse and large amount of data. This pre-trained model can then be fine-tuned on a downstream task, such as activity recognition in the smaller datasets, such as Capture-24. In the main text, Fig. 7 illustrates a multi-task self-supervised approach for feature learning in HAR. We treated each of the tasks as a binary problem predicting whether a self-supervised transformation has been applied. Our multi-task SSL training relied on the *unlabelled UKB*, which contains roughly 700,000 person-days of free-living activity data (100,000 participants, 7 days of wear). For more information we refer the reader to our previous work^26^.

#### B.2.2 Deep Network Architecture

We used a deep convolutional neural network (DCNN) with a ResNet-V2 architecture, consisting of 18 layers and 1D convolutions^57^ as a feature extractor (10M parameters). The learned feature vector was of size 1024. All the tasks will share the same feature extractor. Then, we attached a softmax layer for each of the self-supervised tasks. In the downstream evaluation, we added a fully-connected (FC) layer of size 512 in between the feature extractor and softmax readout. The network structure was fixed for all the downstream evaluations. We computed the cross-entropy loss for each task and weighed all the tasks equally in loss calculation.

### B.3 Hidden Markov Model (HMM) smoothing

Human activity recognition (HAR) model predictions are essentially independent—meaning that the sequence of activities over each 30 second epoch incorporates no temporal information epoch-to-epoch, for instance how the previous epoch prediction affects the current, or next, activity prediction. In order to add temporal dependency to the human activity recognition (HAR) model developed a Hidden Markov Model (HMM) was implemented in a post-processing step to obtain a more accurate sequence of predicted activities over the continuous 14-day data collection period.

**Figure B.II.**
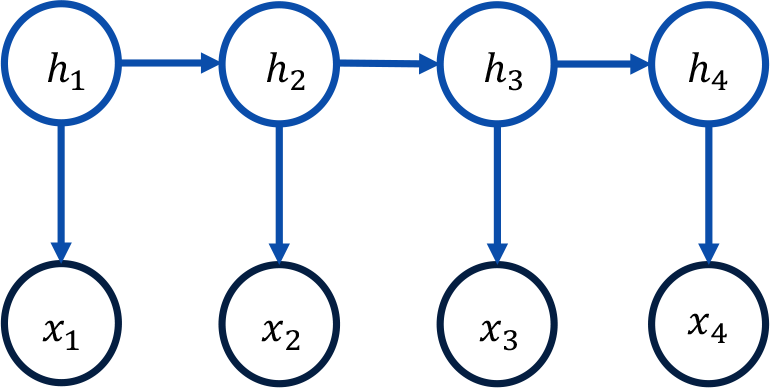
Diagram of a Hidden Markov Model (HMM). The sequence of discrete hidden states **h** = {*h*_1_, …, *h*_(*t*−1)_, *h*_*t*_, *h*_(*t*+1)_, … *h*_*N*_} form a Markov chain. At each time step an observation is obtained by a draw from a probability distribution that is conditional on the value of *h* at that time. This results in a sequence of observable values **x** = {*x*_1_, …, *x*_(*t−*1)_, *x*_*t*_, *x*_(*t*+1)_, …, *x*_*N*_}.

#### Hidden Markov Model (HMM)

The Hidden Markov Model (HMM) defines a Markov chain on hidden (or “latent”) variables *h*_*t*_ = {*h*_1_, *h*_2_, …, *h*_*H*_}, such that only the recent past influences the future:

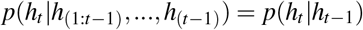

The observed (or “visible”) variables are dependent on the hidden variables through an emission *p*(*x*_*t*_ *h*_*t*_). This defines a joint distribution:

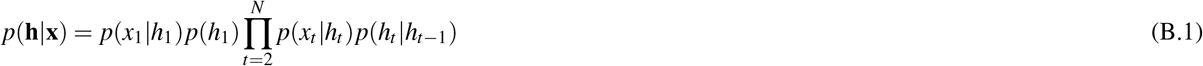

where *p*(*x*_*t*_|*h*_*t*_) defined the emission probability; *p*(*h*_*t*_|*h*_*t−*1_) defines the transition probability between hidden states; The transition distribution *p*(*h*_*t*+1_|*h*_*t*_) is defined by a *H ×H* transition matrix: 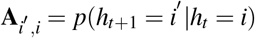. The emission distribution, *p*(*x*_*t*_|*h*_*t*_), has discrete states *x*_*t*_ ∈ {1,…,*V* }, we can define a *V × H* emission matrix: **B**_*i*, *j*_ = *p*(*x*_*t*_ = *i*|*h*_*t*_ = *k*). For continuous outputs, *h*_*t*_ selects one of *H* possible output distributions *p*(*x*_*t*_|*h*_*t*_), *h*_*t*_ ∈ {1,…, *H*}. The most likely hidden path, i.e., sequence of states, 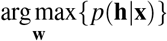, is then found via the is found via the Viterbi algorithm. A diagram of a HMM is shown in Fig. B.II.

At each time step *t*, we define *h*_*t*_ as one of *k* activity classes, *c*_1_, *c*_2_, …, *c*_*k*_ as {sleep, sedentary, light physical activity, moderate-to-vigorous physical activity (MVPA)}. While *h*_*t*_ are not observed directly, at each *t* step there is an dependent observed stochastic emission *x*_*t*_. The hidden state sequence **h** is defined as the true activity labels and the emission distribution *p*(*x*_*t*_ | *h*_*t*_) is estimated directly using the predicted activity probabilities from the HAR model in the training set. As such we use the training predictions of activity from HAR model to infer the most likely sequence of true activity states that would have given rise to those predictions.

**Table B.I.**
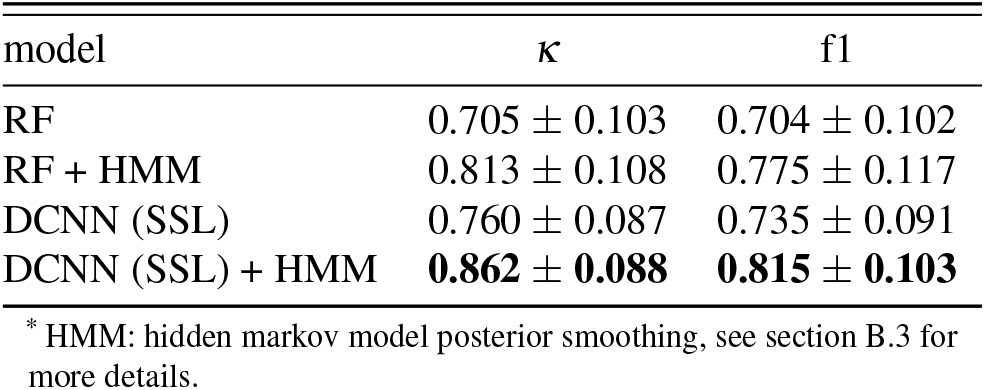
Comparison of activity recognition performance in the Capture-24 dataset between baseline random forest (RF) model and ResNet-based deep convoutional neural network (DCNN), pre-trained on 700,000 person days in the UK Biobank following a self-supervised learning (SSL) framework. *κ*, Cohen’s kappa statistic; F_1_, macro-F1 score.

| model | $\kappa$ | f1 |
| --- | --- | --- |
| RF | $0.705 \pm 0.103$ | $0.704 \pm 0.102$ |
| RF + HMM | $0.813 \pm 0.108$ | $0.775 \pm 0.117$ |
| DCNN (SSL) | $0.760 \pm 0.087$ | $0.735 \pm 0.091$ |
| DCNN (SSL) + HMM | **$0.862 \pm 0.088$** | **$0.815 \pm 0.103$** |

\* HMM: hidden markov model posterior smoothing, see section B.3 for more details.

HMM smoothing helps to correct for erroneous predictions, such as when the transitions between those two classes of activity are rare, for instance sleeping to walking.

**Figure B.III.**
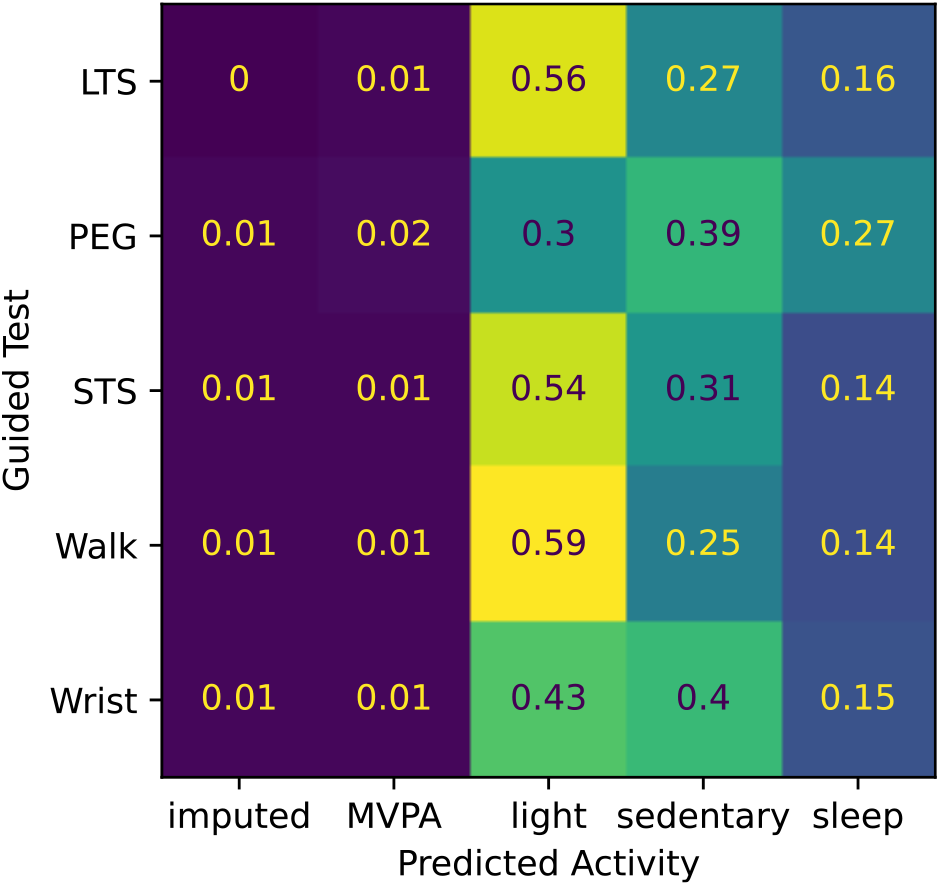
Validation of SSL-HMM activity predictions in the weaRAble-PRO study. Normalised confusion matrix evaluation of the SSL DCNN HAR model on the weaRAble-PRO study with guided test timings as pseudo-labels.

### B.4 Evaluation of activity recognition model

The performance of the SSL model, compared to a feature-based Random Forest (RF) as a baseline, is reported in table B.I. It was observed that the SSL model improved activity recognition performance in Capture-24 beyond feature-based approaches and training a model end-to-end.

Furthermore, an insight into the performance of the SSL DCNN HAR model in the weaRAble-PRO dataset was obtained from a set of simple experiments. While the Capture-24 study uses a similar device (Axivity AX3) and placement (non-dominant wrist) to the weaRAble-PRO’s Apple Watch, the evaluation of the HAR predictions are unknown due to the lack of activity labels in the weaRAble-PRO study. However, as participant’s performed prescribed guided tests during the day, HAR predictions could be benchmarked against the timing of these assessments. As such, guided test timings can act as pseudo-labels in order to evaluate the SSL DCNN HAR model’s robustness in applied to the weaRAble-PRO study, shown in Fig. B.III. As expected, during activity-based guided test assessments, such as walking or sit-to-stand and lie-to-stand, the HAR model more often predicts that participants are performing light activity. Guided assessments that require participants to be stationary while performing the task, such as the PEG test or wrist ROM test, are predicted more as sedentary activities. The percentage of sleep-based predictions during the guided test assessments (although incorrect) are roughly in line with the overall probability of daytime sleep, as observed depicted in figures A.Ic–A.Id. Further work is needed to fully characterise and appraise the predictions of daytime sleep, which are assumed to be incorrect predictions of sedentary activity.

### B.5 Passive Features

The section below details the passively extracted, activity monitoring features in the weaRAble-PRO study. Activity monitoring features were developed based on broad activity prediction labels {sleep, sedentary, light physical activity, moderate-to-vigorous physical activity (MVPA)}^28,29^ and fine-grained activity prediction labels {sleep, sitting/standing, mixed, vehicle, walking, bicycling}^27^

Measures of physical activity and sleep were summarised based on smartwatch actigraphy sensor data magnitude data per epoch, or aggregated by intensity levels, activity classification and bouts of activity. These features could be broadly grouped into physical activity domains: total volume of daytime activity (TVDA), fragmentation of activity (AF), along with two RA symptom-specific domains focusing on morning stiffness (MORN) and night-time restlessness (NTR):

1. **Total volume of daytime activity (TVDA)**: captures information around the overall physical activity and the ability to perform physical activity at varying levels of intensity during daytime, which are known to be altered in patients with RA^13, 58^;
2. **Activity Fragmentation (AF)**: Metrics in this domain attempt to capture information related to the ability to perform sustained activity. Frequent interruptions of physical activity may reflect a worse health condition. For example, RA patients may need to interrupt some activity due to increased joint pain.
3. **Morning stiffness (MORN)** is a common symptom of RA. These measures include information related to timing of activity after getting up in the morning as estimated from the activity classification^1^.
4. **Night-time Restlessness (NTR)**: These measures include information related to timing, duration and quality of sleep. There is evidence that RA patients experience fluctuations in disease activity following a circadian rhythm with worsening of the illness during the night^59^, thus measures of overnight movement serve as a proxy for estimating night-time restlessness, reflecting the impact of the disease on sleep quality.

## C Methodology: Smartphone sensor feature extraction

### C.1 Smartphone guided tests administered in weaRAble-PRO

**Table C.I.** Overview of the daily prescribed smartphone-recorded Active Assessments (denoted “Guided Tests”) in the weaRAble-PRO study.

| Exercise | Abbrev. | Brief Description |
| --- | --- | --- |
| Wrist ROM | WRT | Participants were requested to sit down and place their forearm at the edge of a table, holding the iPhone horizontally facing up in their hand, and to flex and extend their wrist joint to its maximum angle, repeating the motion for 10 seconds. The test was carried out twice, once using each hand<sup>12</sup>. |
| Gait | WLK | Participants were asked to affix the iPhone to their leg (on the right thigh) facing outward using a provided strap, then to walk in a straight line for 30 seconds (while being allowed to turn around at any point in the middle of the test)<sup>12</sup>. |
| 9-hole peg test | PEG | Digital touch-screen version of the standard clinical assessment where participants are asked to place, and subsequently remove, 9 pegs into and from a round hole, in the fastest time possible<sup>33</sup>. |
| Sit-to-stand | STS | Participants were requested to perform a sit-to-stand transition from a chair with the iPhone attached to their upper right thigh with a strap, repeating the exercise 5 times at their own pace<sup>30,34</sup>. |
| Lie-to-stand | LTS | Participants performed a lie-to-stand transition: from lying still with legs stretched on a bed, to standing up on the floor. Participants were requested to affix the iPhone to their right thigh with a strap during the exercise and repeat twice at their own pace<sup>30,34</sup>. |

**Abbreviations** (abbrev.): ROM, range of motion;

### C.2 Peg Test Algorithm

The 9-hole peg test is a two-step test of hand dexterity to measure the MSFC score in Multiple Sclerosis, or signs of Parkinson’s disease or stroke^60^. The digital 9-hole peg test (9HPT), and subsequent metrics calculated, are proprietary to Apple ResearchKit, see http://researchkit.org/docs/docs/ActiveTasks/ActiveTasks.html for more details. This smartphone version of the 9HPT task requires participants to use two fingers to touch the on-screen peg and drag it into the an on-screen hole. Once all 9 pegs have been placed in each hole, the task is repeated by removing the pegs in the same manner. The 9HPT is repeated using both dominant and non-dominant hands. The total duration that the user spent taking the test for each hand repetition is recorded.

### C.3 Wrist ROM Test Algorithm

The wrist range of motion (ROM) test algorithm is outlined previously as part of the GSK PARADE study^11,12^. The iPhone accelerometer sensor data is converted to angular positions; ROM is then computed based on the differences between angular maxima; angular velocity is determined from the gyroscope sensors.

### C.4 Walk Test Algorithm

The gait test algorithm is outlined previously as part of the GSK PARADE study^12^. During walking, initial (IC) and final (FC) feet contact timings are calculated from the smartphone accelerometer data with an inverted pendulum model, as described in^61,62^. Contact points are determined by integration of the vertical component of the accelerometer signal, **a**_*y*_, and subsequent differentiation of that signal with a continuous wavelet transform (CWT, convolution of the accelerometer data and an analysing function, i.e., mother wavelet). ICs and FCs are then denoted by minima and maxima timings respectively in this transformed signal. Detected peaks were used to estimate the number of steps, cadence, step length^63^, and walk velocity for each assessment.

### C.5 Sit-to-stand Test Algorithm

Both the iPhone accelerometer and gyroscope data were used to determined when participants were in a sitting or standing position during the sit-to-stand (STS) exercise^30^. The vertical accelerometer component, **a**_*y*_, determined sit-to-stand transitions by estimating the phone axis orientation (and change thereof) relative to gravity, as the participants moved between sitting and standing—given the phone’s axis should be fixed as it is strapped to the participants thigh using a strap. Gyroscope axis sensor components, (**g**_*x*_, **g**_*y*_, **g**_*z*_), helped determine whether a participant had completed valid transitions during the exercise. The start and end points of the STS transitions could therefore be determined by identifying peaks in **a**_*y*_ that were within the given thresholds where participants could be feasibly standing or lying. The length of a sit-to-stand transition was then calculated as the difference between the point where the participant first reached a standing position and the final point where the participant was sitting prior to beginning a standing motion.

### C.6 Lie-to-Stand Test Algorithm

Following a similar STS algorithm^30^, during the lie-to-stand test, accelerometer and gyroscope (y-axis) measurements were used to determine a participant’s standing and lying transition points following the algorithm introduced in^64^. Gyroscopic y-axis gravity relates the phone’s orientation relative to gravity at a given moment in time and the phone acceleration helped to improve accuracy in determining participants’ lying positions—for example, the point of minimum acceleration generally corresponded to moments when participants held a lying position (causing a short plateau in y-gravity).

## D Further Study Details

### D.1 Inclusion and Exclusion Criteria

For participation in the full pilot study, 30 RA participants and 30 HCs matched on age, gender, and race were recruited. Due to the small sample size and potentially limited pool for recruitment, ages were matched within a window of 3 ± years.

The overall ratio of moderate to severe participants was chosen not to exceed 2:1 in either direction. Inclusion/Exclusion criteria were the same for RA participants and HCs, unless otherwise noted. All participants must have been able and willing to perform the pre-defined guided tests at the start of the study, follow audio instructions from an iPhone, and have a sufficient level of English language to ensure ability to understand mobile app and questionnaires.

Rheumatoid Arthritis participants, at least 21 years of age at date of consent for study, were selected based on clinically verified diagnosis of moderate-to-severe RA, with severity assessed using Routine Assessment of Patient Index Data 3 (RAPID3). Healthy controls were selected based on no prior or current diagnosis of a rheumatological disorder, inflammatory disorder, malignancy, or other relevant diseases. Further exclusion criteria for all participants included history of other inflammatory rheumatologic or systemic autoimmune disorder (e.g., Hashimoto’s thyroiditis or Sjogren’s syndrome), history of movement disorders, other neurological disorders or conditions resulting in significant physical impairments that impact joint movements to be assessed, history of postural hypotension, unexplained syncope, or other conditions that make it difficult for participants to perform guided tests such as the lie-to-stand test and any history of severe skin allergy. Participants were also excluded if they required use of a wheelchair, walking aids, artificial limbs, or had any active implantable device, such as a pacemaker.

### D.2 Data collection timelines

**Figure D.I.**
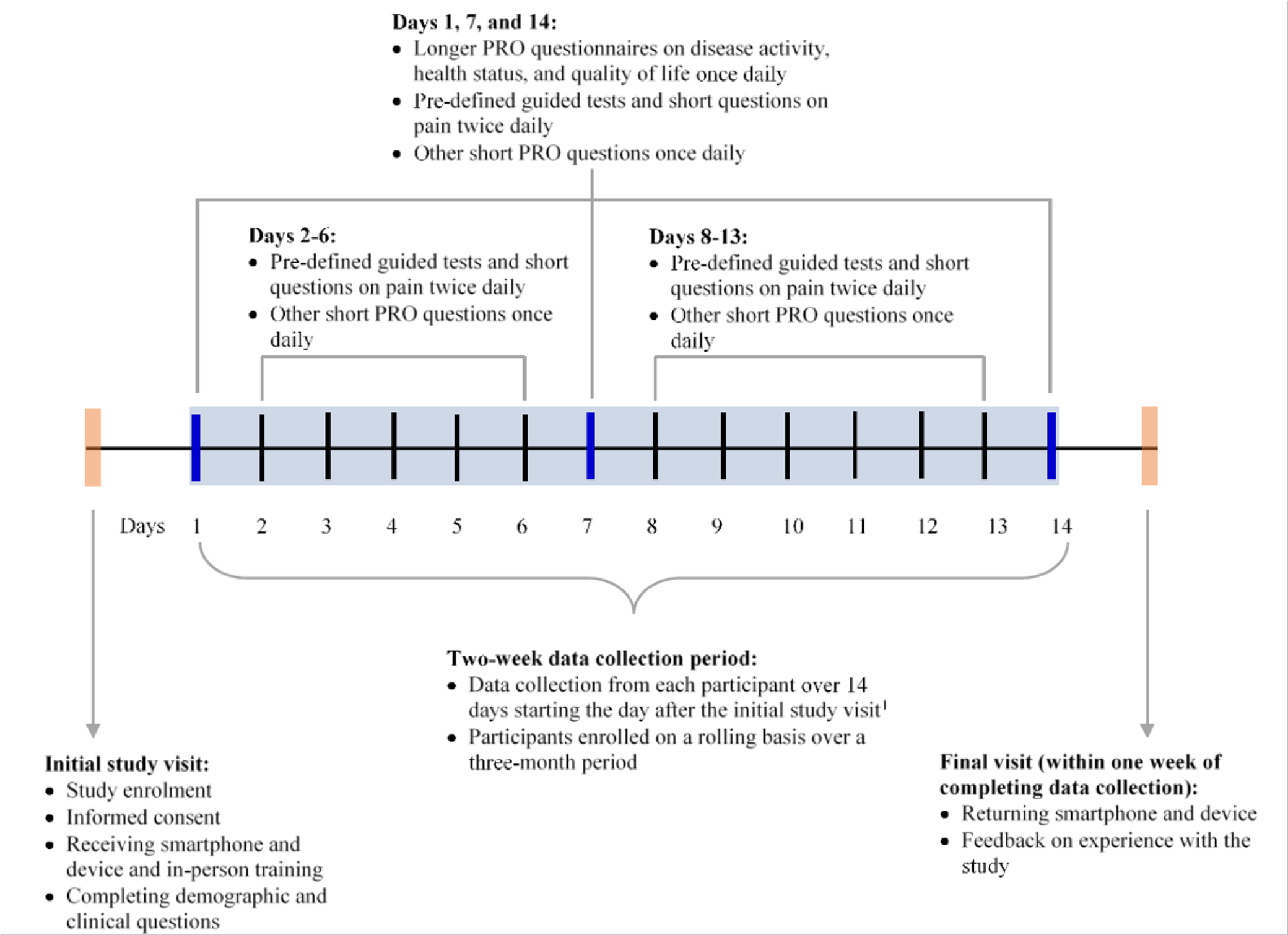
Overview of frequency and timeline of data capture for each data source in the weaRAble-PRO Study.

### D.3 Patient-reported outcomes administered in weaRAble-PRO

**Table D.I.**
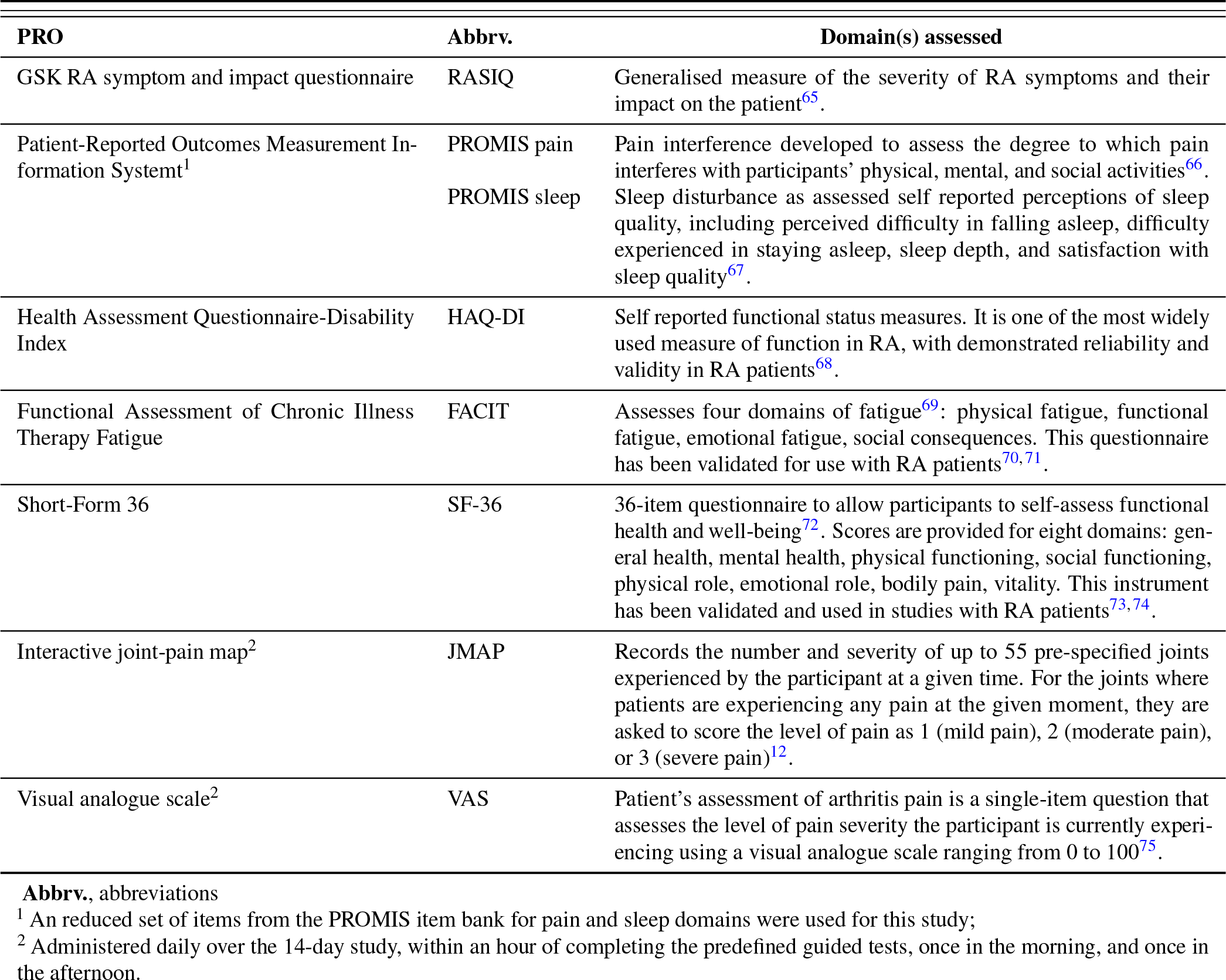
Overview of the PRO assessments administered to RA and HC participants on days 1, 7, and 14 in the weaRAble-PRO study

## E. List of Extracted Features

**Table E.I.**
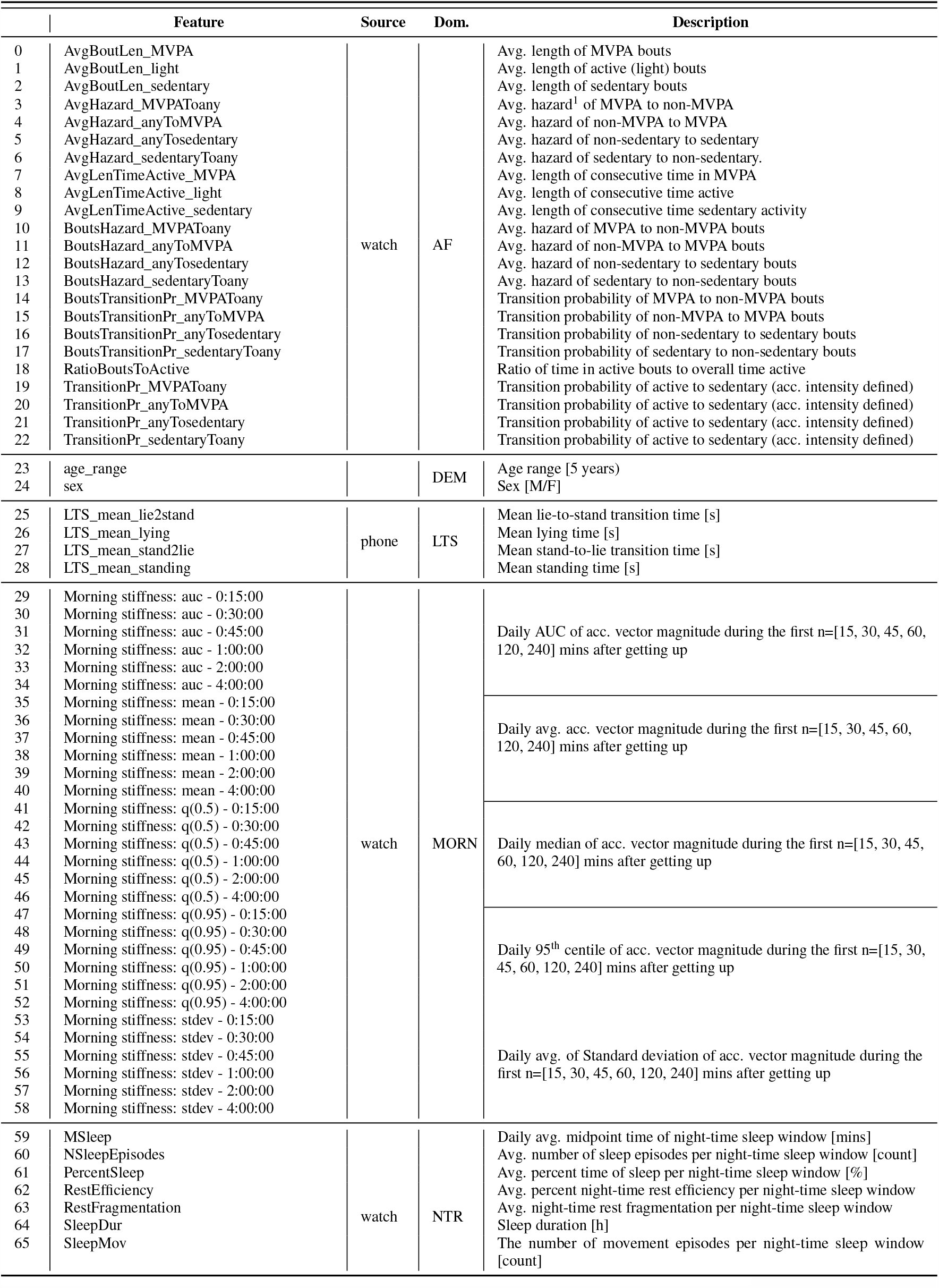
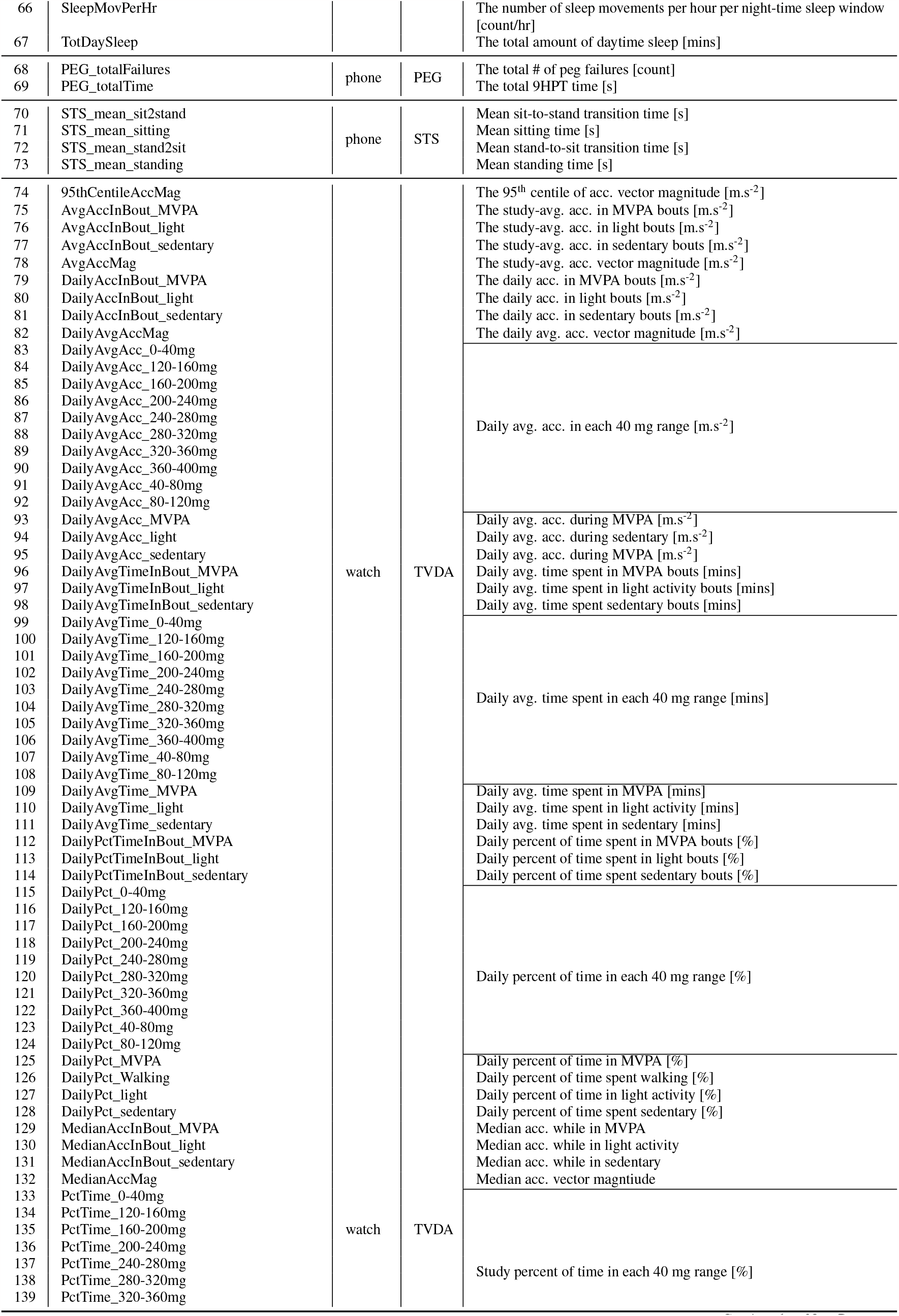
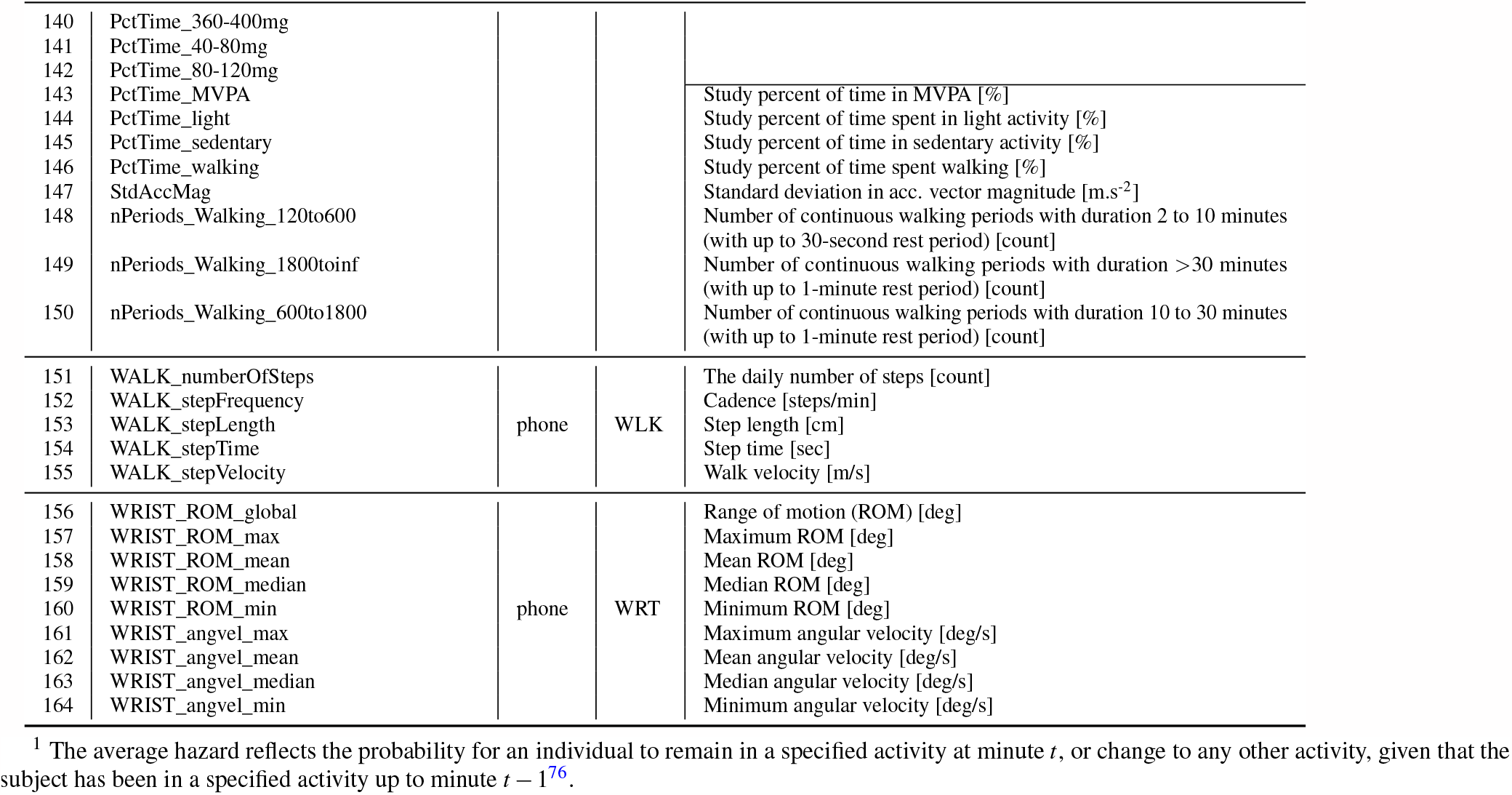
Description of the sensor-based features extracted in the weaRAble-PRO study

## F Methodology: Machine learning analysis for characterising RA

### F.1 Feature Pre-processing

Missing data extracted from the GTs and passive monitoring was imputed using a carry-last value forward per participant^77^. In cases where the last value was missing, mean imputation was used instead.

Features were assessed for non-normality by visual inspection. Those non-normal features were transformed using box-cox transformations^78^.

Features were normalised using the z-score to have unit variance using their respective mean *μ* and standard deviation 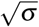.

### F.2 Classification and Regression Models

#### F.2.1 Linear Regression

A regression model explicitly describes a relationship between predictor(s) **X** ∈ ℝ ^*N×P*^ and continuous response variables **y** ∈ ℝ^*N*^, the most basic of which is *linear regression* (LR)^55^. For an *i*^*th*^ observation row of **X, x** *≡* **x**_**i**_ ∈ ℝ^1*×P*^:

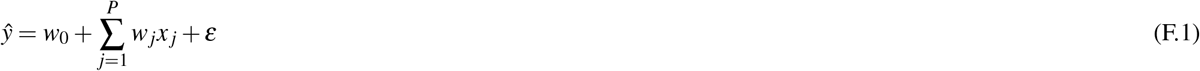

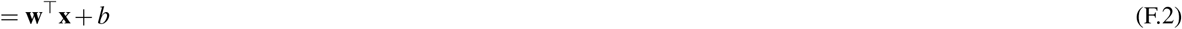

where *w*_*j*_ values denote the slope (weights, or regression coefficients) of the *x* _*j*_ features; *w*_0_ is the intercept term; and *ε* denote the residual (model) errors term, which are assumed to be normally distributed with constant variance, *ε ∼ 𝒩* (0, *σ* ^2^)^48,55^. Often a linear model is described in vector notation (F.2), **w** = [*w*_0_, *w*_1_, *w*_2_, …, *w*_*P*_], where *w*_0_ is denoted as the “bias”, *b* term, and the *ε*-term is often omitted.

#### F.2.2 Logistic Regression

Generalised linear models (GLMs) are extensions of linear regression models that can have non-linear outputs^79^. GLMs utilise canonical link functions, *φ*, to transform the outputs of a linear regression: *ϕ* = **w**^*⊤*^**x** to another distribution, such as with a logistic *φ* = *σ* (*ϕ*) link function (or inversely the logit, representing the log-odds) which will be used to form Logistic Regression for binary classification tasks^48,80^, in this case *φ* is sigmoidal and is bounded between [0, 1], therefore the output of *σ* can be interpreted as the probability of *y* = 1:

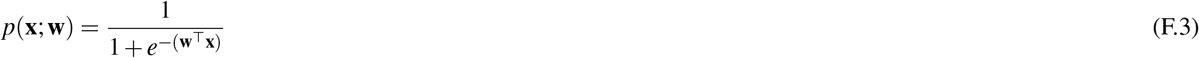

A threshold can be applied to the probabilistic output *p* to determine a classification prediction 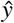 for a Logistic Regression model; threshold values are typically chosen as 0.5, but this can be altered based on the use case.

##### Regularisation as Feature Selection

Many statistical and machine learning models can easily overfit to the training data, resulting in poorer estimations, models that are not generalisable or too complex. This is likely in the weaRAble-PRO dataset where we have *P≫ N* problem: the number of predictors *P* is much larger than the number of observations *N* (participants), a case where standard models fail.

Regularisation can be introduced to mitigate against the *p ≫n* problem. For example large coefficient values in a regression can be penalised by adding a regularisation term to a loss function, or through reducing the number of parameters or features used in a model. The most common regularisers use the *𝓁*_*p*_-norm defined by^48,81^:

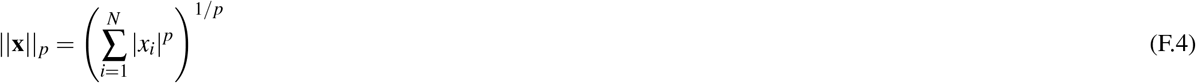

for any **x** ∈ ℝ^*N×*1^, where the real number *p ≥*1 defines the *𝓁*_*p*_ space. In this work, we experimented with a number of regularisation techniques in order to perform the classification tasks introduced in section 2.2.

#### F.2.3 Least Absolute Shrinkage and Selection Operator

The Least Absolute Shrinkage and Selection Operator (LASSO)^81,82^ is a technique that conversely solves the *𝓁*_1_ − penalised sum of squares (F.4) in a linear regression such that:

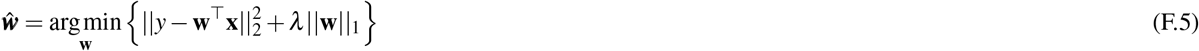

This is equivalent to minimising the sum of squares with a constraint of the form: 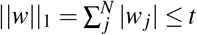. Because of the form of the *𝓁*_1_-penalty, LASSO both shrinks coefficients but also encourages sparsity in a model’s parameters and thus inherently forms feature selection, shrinking non-important features to zero. The LASSO can also be extended to perform feature selection for classification by substituting a canonical link function (such as the logistic *φ* = *σ* (*ϕ*)) and following the same procedure outlined in equation F.3, essentially performing regularised-logistic regression (denoted LR-lasso in this work)^81^.

#### F.2.4 Ridge Regression

Ridge Regression (Tikhonov Regularisaion)^48,81^ is a technique which utilises the *𝓁*_2_-norm to impose a penalty on the size of the coefficients in a linear regression (F.4) such that:

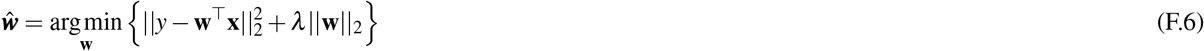

This is equivalent to minimising the sum of squares with a constraint of the form: 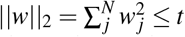 (where *t* controls the amounts of shrinkage; there is an exact relationship between *t* and corresponding *λ* (denoted LR-ridge in this work).

#### F.2.5 Elastic Net

Elastic net linearly combines the *𝓁*_1_ and *𝓁*_2_ penalties of the lasso and ridge methods such that:

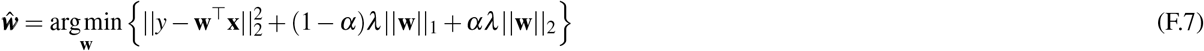

given non-negative values *λ*, and *α* that is strictly between 0 and 1, which determines the trade off between *𝓁*_1_ and *𝓁*_2_ regularisation (denoted LR-elastic-net in this work).

#### F.2.6 Sparse-Group LASSO

The sparse-group lasso is an extension of the lasso that promotes both group sparsity and within group parameter-wise sparsity, through a group lasso penalty and the lasso penalty:

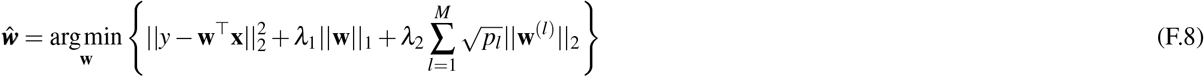

where *λ*_1_ is the parameter-wise regularisation penalty and *λ*_2_ is the group-wise regularisation penalty. The data **x** contains sub-grouping, such that **x**^(*l*)^, denoting the features in group *l*, and corresponding learned weights **w** containing contains sub-grouping **w**^(*l*)^; *p*_*l*_ is the length of **w**^(*l*)^ (i.e., the number of features in each group) and *M* are the total number groups. Therefore, the sparse group lasso penalty will yield a sparse set of groups and also a sparse set of covariates in each selected group. As denoted in^50^, we use the term “groupwise sparsity” to refer to the number of groups with at least one nonzero coefficient, and “within group sparsity” to refer to the number of nonzero coefficients within each nonzero group (denoted LR-SG-lasso in this work).

#### F.2.7 Random Forest

Classification and Regression Trees (CART), specifically Random Forests (RF), are a multi-functional, non-linear method capable of performing regression, classification and feature selection^51^. Unlike the linear filter-based methods of feature selection, for example, lasso, RFs incorporate non-linear feature selection as part of the model methodology.

Random Forests consist of a large ensemble of decision trees arranged in a hierarchical structure. To build an individual tree, we recursively descend through the hierarchy, performing binary splits (decisions) at each level in the structure (a node, *j*) using a single feature *x* _*j*_ *∈ χ* ^*p*^ based on a threshold value (splitting criterion) *s* _*j*_, sub-partitioning the feature space χ_*j*_ at each node. A tree is typically expanded until all leaves are pure (i.e each partition χ _*j*_ represents only one class) or until all leaves contain less than the minimum number of samples in a partition χ _*j*_ required to split a node. An individual tree selects *m < N* random subset of observations (with replacement), and each node considers a random subset of *p* features for each split. Once all trees have been grown, each of the ‘weaker’ decisions are aggregated (or *ensembled*) creating a robust final prediction. For example, consider an ensemble of two trees. The prediction scores of each individual tree are summed to obtain the final prediction:

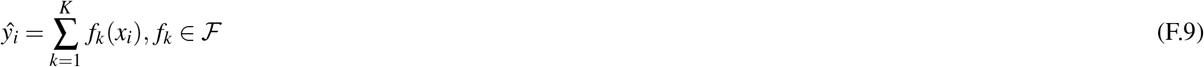

where *K* is the number of trees, *f*_*k*_ is a function in the functional space ℱ, and ℱ is the set of all possible CARTs. The objective function optimised is then given by:

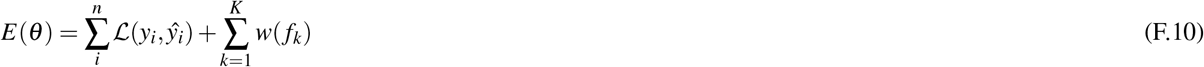

where *w*(*f*_*k*_) is the complexity of the tree *f*_*k*_.

Classification predictions are deduced as the majority class label of the observations present in each final partition χ_*j*_, whereas for continuous prediction (i.e., regression) the mean of the (continuous) responses would be calculated instead. To determine the optimal split criterion *s* _*j*_ for each *x* _*j*_ to create each χ _*j*_ we evaluate the *Gini* importance, which quantifies the average gain of purity (i.e., the presence of one class) caused by splits of a given variable. For regression the mean decrease in mean square error (MSE) is assessed instead.

#### F.2.8 Extreme Gradient Boosted Trees (XGB)

Extreme Gradient Boosted Trees (XGB) iterates on the CART through a regularising gradient boosting framework—the difference being in how trees are built and combined. Rather than bagging, like in CART, specifically RF, gradient boosting improves a single weak model by combining it with a number of other weak models in order to generate a collectively stronger model^52^. This boosting is formed as additive strategy during training where a gradient descent algorithm is used to minimise (or maximise) an objective function *E*(*θ*) for each new tree that is added at each time step, *t*:

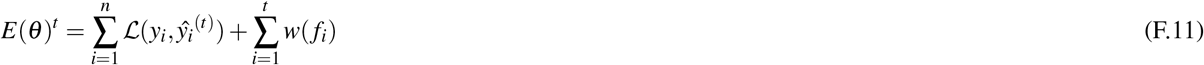

where 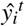 is the prediction value at step *t*; and *f*_*i*_ are the parameters of a tree, i.e., the tree structure and the leaf scores that are needed to be learned; and *w*(*f*_*i*_) is the complexity of the tree. Therefore 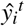 is determined by the prediction of the previous tree at *t −* 1, 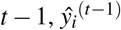:

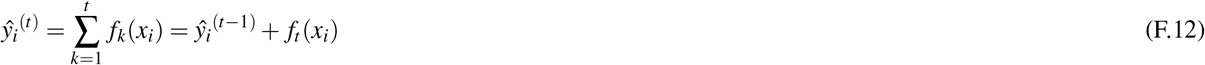

To optimise the error function, XGBoost computes gradients (Jacobian) and Hessians of the error, denoted below as:

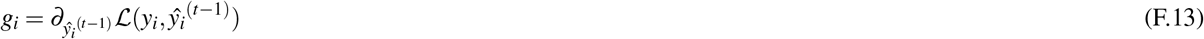

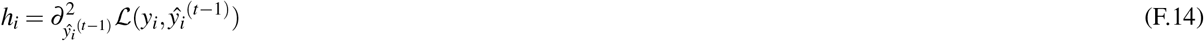

which can be subbed into the objective function in equation (F.11), yielding:

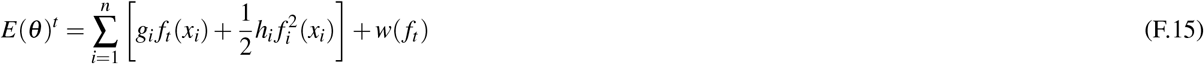

Next, to define the complexity of the tree *w*(*f*), we first refine the function for the definition of a tree *f* (*x*) as:

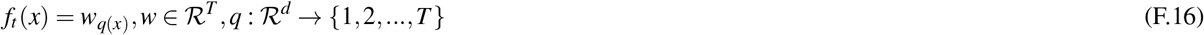

where *w* is the vector of scores on leaves, *q* is a function assigning each data point to the corresponding leaf, and *T* is the number of leaves. Then, in XGBoost, we can define the complexity over the number of leaves, *T* :

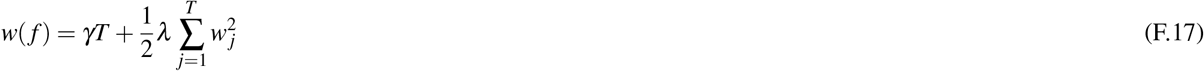

where *γ* and *λ* are tune-able regularisation parameters. Subbing equation F.17 into equation F.15, becomes:

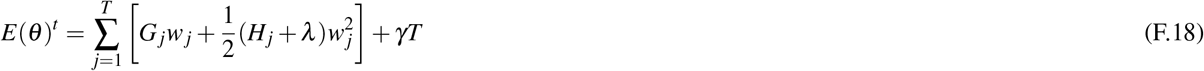

where 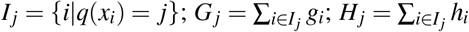. Following the convention in^52^, equation F.18 can be reformulated as solving over:

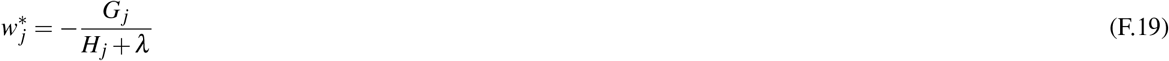

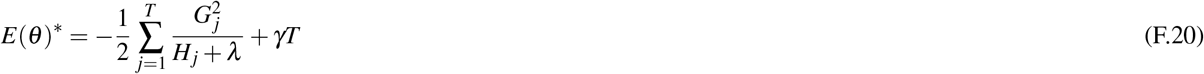

Summarising, XGB iteratively trains an ensemble of shallow decision trees, with each iteration using the error residuals of the previous model to fit the next model. The objective *E*(*θ*)^*\**^ measures how good a tree structure *q*(*x*) is, optimising one level of the tree at a time, while regularising on the model complexity. For example, to split a leaf into two leaves, and the score will be:

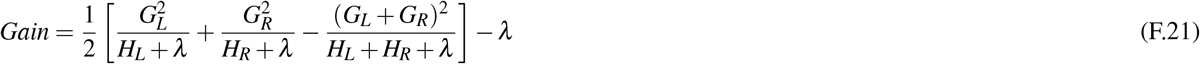

if the gain acquired by adding a branch is smaller than *γ*, it would not be added.

### F.3 Parameter Tuning

Optimal model parameters were determined via randomised grid-search over internal 5-fold (subject-wise) CV with 500 iterations. In the case of XGB, early stopping was determined using roughly 10% of the training data, proportionally, as validation, with for 10 boosting rounds.

For regularised logistic regression models, a parameter search was determined over *𝓁*_1_ and *𝓁*_2_ regularisation terms on weights, *λ* ∈ {10^*−*5^, …, 10^*−*1^, …, 0, …, 10^0^, 10^1^, …, 10^5^}; and the elastic-net mixing parameter *α ∈ {*0, 0.1, …, 1};

RFs have relatively little hyperparameter tuning: the number of trees to build *p ∈ {*500, 1000, 1500}; the number of input variables chosen at each node 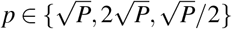, where *P* are the number of features, as suggested in^83,84^.

For XGB, a parameter search was determined over: the boosting learning rate *p ∈* {0.01, 0.05, 0.1, 1} ; number of boosting rounds *p ∈*{ 100, 500, 1000, 1500 }; maximum tree depth for base learners *p ∈*{ 3, 4, 5, 8, 10 }; the subsample ratio of the training instance (selecting random training instances with higher probability when the gradient and hessian are larger) *p ∈ {*0, 0.5, 1}; the subsample ratio of features when constructing each tree *p ∈ {*0.2, 0.6, 0.8, 1.0}; minimum sum of instance weight (hessian) needed in a child *p* ∈ {1, 5, 10, 50, 100}; the *𝓁*_1_ and *𝓁*_1_ regularisation term on weights, *p ∈ {*10^*−*5^, …, 10^*−*1^, …, 0, …, 10^0^, 10^1^, …, 10^5^}

